# Psychiatric disorders converge on common pathways but diverge in cellular context, spatial distribution, and directionality of genetic effects

**DOI:** 10.1101/2025.07.11.25331381

**Authors:** Worrawat Engchuan, Omar Shanta, Kuldeep Kumar, Jeffrey R. MacDonald, Bhooma Thiruvahindrapuram, Omar Hamdan, Marieke Klein, Adam Maihofer, James Guevara, Oanh Hong, Guillaume Huguet, Molly Sacks, Mohammad Ahangari, Rayssa M.M.W. Feitosa, Kara Han, Marla Mendes, Xiaopu Zhou, Nelson X. Bautista, Giovanna Pellecchia, Zhouzhi Wang, Daniele Merico, Ryan K.C. Yuen, Brett Trost, Ida Sønderby, Mark J. Adams, Rolf Adolfsson, Ingrid Agartz, Allison E. Aiello, Martin Alda, Judith Allardyce, Ananda B. Amstadter, Till F.M. Andlauer, Ole A. Andreassen, María S. Artigas, S. Bryn Austin, Muhammad Ayub, Dewleen G. Baker, Nick Bass, Bernhard T. Baune, Maximilian Bayas, Klaus Berger, Joanna M. Biernacka, Tim Bigdeli, Jonathan I. Bisson, Douglas Blackwood, Marco Boks, David Braff, Elvira Bramon, Gerome Breen, Tanja Brueckl, Richard A. Bryant, Cynthia M. Bulik, Joseph Buxbaum, Murray J. Cairns, Jose M. Caldas-de-Almeida, Megan Campbell, Dominique Campion, Vaughan J. Carr, Enrique Castelao, Boris Chaumette, Sven Cichon, David Cohen, Aiden Corvin, Nicholas Craddock, Jennifer Crosbie, Darrina Czamara, Udo Dannlowski, Franziska Degenhardt, Douglas L. Delahanty, Astrid Dempfle, Guillaume Desachy, Arianna Di Florio, Faith B. Dickerson, Srdjan Djurovic, Katharina Domschke, Lisa Douglas, Ole K. Drange, Laramie E. Duncan, Howard J. Edenberg, Tonu Esko, Steve Faraone, Norah C. Feeny, Andreas J. Forstner, Barbara Franke, Mark Frye, Dong-jing Fu, Janice M. Fullerton, Anna Gareeva, Linda Garvert, Justine M. Gatt, Pablo Gejman, Daniel H. Geschwind, Ina Giegling, Stephen J. Glatt, Joe Glessner, Fernando S. Goes, Katherine Gordon-Smith, Hans Grabe, Melissa J. Green, Michael F. Green, Tiffany Greenwood, Maria Grigoroiu-Serbanescu, Raquel E. Gur, Ruben C. Gur, Jose Guzman-Parra, Jan Haavik, Tim Hahn, Hakon Hakonarson, Joachim Hallmayer, Marian L. Hamshere, Annette M. Hartmann, Arsalan Hassan, Caroline Hayward, Johannes Hebebrand, Sian M.J. Hemmings, Stefan Herms, Marisol Herrera-Rivero, Anke Hinney, Georg Homuth, Andrés Ingason, Lucas T. Ito, Nakao Iwata, Ian Jones, Lisa A. Jones, Lina Jonsson, Erik G. Jönsson, René S. Kahn, Robert Karlsson, Milissa L. Kaufman, John R. Kelsoe, James L. Kennedy, Anthony King, Tilo Kircher, George Kirov, Per Knappskog, James A. Knowles, Nene Kobayashi, Karestan C. Koenen, Bettina Konte, Mayuresh Korgaonkar, Kaarina Kowalec, Marie-Odile Krebs, Mikael Landén, Claudine Laurent-Levinson, Lauren A. Lebois, Doug Levinson, Cathryn Lewis, Qingqin Li, Israel Liberzon, Greg Light, Sandra K. Loo, Yi Lu, Susanne Lucae, Charles Marmar, Nicholas G. Martin, Fermin Mayoral, Andrew M. McIntosh, Katie A. McLaughlin, Samuel A. McLean, Andrew McQuillin, Sarah E. Medland, Andreas Meyer-Lindenberg, Vihra Milanova, Philip B. Mitchell, Esther Molina, Bryan Mowry, Bertram Muller-Myhsok, Niamh Mullins, Robin Murray, Markus M. Nöthen, John I. Nurnberger, Kevin S. O’Connell, Roel A. Ophoff, Holly K. Orcutt, Michael J. Owen, Aarno Palotie, Carlos Pato, Michele Pato, Joanna Pawlak, Triinu Peters, Tracey L. Petryshen, Giorgio Pistis, James B. Potash, John Powell, Martin Preisig, Digby Quested, Josep A. Ramos-Quiroga, Andreas Reif, Kerry J. Ressler, Marta Ribasés, Marcella Rietschel, Victoria B. Risbrough, Margarita Rivera, Alex O. Rothbaum, Barbara O. Rothbaum, Dan Rujescu, Takeo Saito, Alan R. Sanders, Russell J. Schachar, Peter R. Schofield, Eva C. Schulte, Thomas G. Schulze, Laura J. Scott, Soraya Seedat, Christina Sheerin, Jianxin Shi, Pamela Sklar, Susan Smalley, Olav B. Smeland, Jordan W. Smoller, Edmund Sonuga-Barke, David St. Clair, Nils Eiel Steen, Dan Stein, Frederike Stein, Murray B. Stein, Fabian Streit, Neal Swerdlow, Florence Thibaut, Johan H. Thygesen, Ilgiz Timerbulatov, Claudio Toma, Edward Trapido, Micheline Tremblay, Ming T. Tsuang, Monica Uddin, Marquis P. Vawter, John B. Vincent, Henry Völzke, James T. Walters, Cynthia S. Weickert, Lauren A. Weiss, Myrna M. Weissman, Thomas Werge, Stephanie H. Witt, Miguel Xavier, Robert Yolken, Ross M. Young, Tetyana Zayats, Lori A. Zoellner, AGP Consortium, PEIC Psychosis Endophenotypes International Consortium, ADHD Working Group of the Psychiatric Genomics Consortium, Autism Working Group of the Psychiatric Genomics Consortium, Bipolar Disorder Working Group of the Psychiatric Genomics Consortium, Major Depressive Disorder Working Group of the Psychiatric Genomics Consortium, PTSD Working Group of the Psychiatric Genomics Consortium, Schizophrenia Working Group of the Psychiatric Genomics Consortium, CNV Working Group of the Psychiatric Genomics Consortium, Kimberley Kendall, Brien Riley, Naomi R. Wray, Michael C. O’Donovan, Patrick F. Sullivan, Sandra Sanchez-Roige, Caroline M. Nievergelt, Sébastien Jacquemont, Stephen W. Scherer, Jonathan Sebat

## Abstract

Psychiatric conditions share common genes, but mechanisms that differentiate diagnoses remain unclear. We present a multidimensional framework for functional analysis of rare copy number variants (CNVs) across 6 diagnostic categories, including schizophrenia (SCZ), autism (ASD), bipolar disorder (BD), depression (MDD), PTSD, and ADHD (N = 574,965). Using gene-set burden analysis (GSBA), we tested duplication (DUP) and deletion (DEL) burden across 2,645 functional gene sets defined by the intersections of pathways, cell types, and cortical regions. While diagnoses converge on shared pathways, mixed-effects modeling revealed divergence of pathway effects by cell type, brain region, and gene dosage. Factor analysis identified latent dimensions aligned with clinical axes. A primary factor (F1) captured reciprocal dose-dependent effects of DUP and DEL in SCZ reflecting positive and negative effects in excitatory versus inhibitory neurons and association versus sensory cortex. SCZ and ASD were both strongly aligned with F1 but with opposing directionalities. Orthogonal factors highlighted neuronal versus non-neuronal effects in mood disorders (F2) and differential spatial distributions of DEL effects in ADHD and MDD (F3). High-impact CNVs at 16p11.2 and 22q11.2 were enriched for combinations of cell-type-specific genes involved in pathways consistent with our broader findings. These results reveal molecular and cellular mechanisms that are broadly shared across psychiatric traits but differ between diagnostic categories in context and directionality.

## Background

Genes that are associated with psychiatric conditions carry rich information about the timing, location, and nature of the biological processes that contribute to psychopathology ^1,2^. The molecular functions of genes point to the cellular pathways and regulatory networks that underlie vulnerability to psychiatric disorders. Furthermore, because gene expression is tightly regulated in a cell-type and region-specific manner across the brain, the discovery of genes can also provide insight into the neuroanatomical circuits that influence psychiatric traits. The discovery of hundreds of genes and copy number variations (CNVs) that underlie major psychiatric conditions such as schizophrenia (SCZ)^3–6^ and autism spectrum disorder (ASD)^7–11^ has implicated a variety of pathways including synaptic function, chromatin regulation, cell signaling, cytoskeletal proteins, and DNA and RNA binding proteins that regulate neurodevelopment ^3,12–18^. Similar pathways have been implicated by transcriptome characterization of post-mortem brains from case samples of idiopathic ASD, SCZ and bipolar disorder (BD) ^19–23^. Genes implicated in psychiatric diagnoses are also enriched in specific neural cell types. RNA sequencing in postmortem samples have identified neuronal and glial signatures associated with ASD ^24,25^ and differences in the distributions of glial and neuronal cells in mood disorders ^26^. Analysis of GWAS associations has found enrichment of SCZ ^27^, major depressive disorder (MDD) and post-traumatic stress disorder (PTSD) ^28^ associations in mature excitatory and inhibitory neurons.

Despite significant progress in identifying risk genes and pathways in psychiatric conditions, there remains a limited understanding of how neural processes relate to specific psychiatric traits or diagnoses. Many of the same biological pathways, such as those described above, have been repeatedly associated with multiple diagnostic categories, including SCZ^5,15,29^, BD^14,30^, ASD^16^, intellectual disability ^31^ and congenital heart disease ^32^. Thus, functional convergence that is evident from pathway enrichment analysis of the associated genes highlights broad biological themes but lacks the resolution to differentiate neural mechanisms that differ between diagnostic categories.

CNVs have been shown to exert dose-dependent effects on a range of complex traits, including gene expression^33^, head size ^4,34^, brain volume ^35,36^, functional connectivity^37^, body mass ^38^, craniofacial morphology^39^. As described in our companion paper ^40^, this pattern extends to psychiatric traits, where reciprocal duplications (DUPs) and deletions (DELs) of genes show dose-dependent effects and diverge in their genotype-phenotype associations. A more detailed functional analysis of gene-dosage effects could clarify how alterations in molecular pathways contribute to psychiatric traits. In this study, we developed and applied an integrated framework to examine how gene-dosage effects on pathways, cell types, and brain regions relate to clinical diagnoses (**Fig. 1**). Key elements of this approach include accounting for (1) directionality of gene-dosage effects and their distribution within (2) neural cell-types and (3) cortical brain regions, and we perform a comparative analysis across multiple diagnostic categories.

**Fig. 1:**
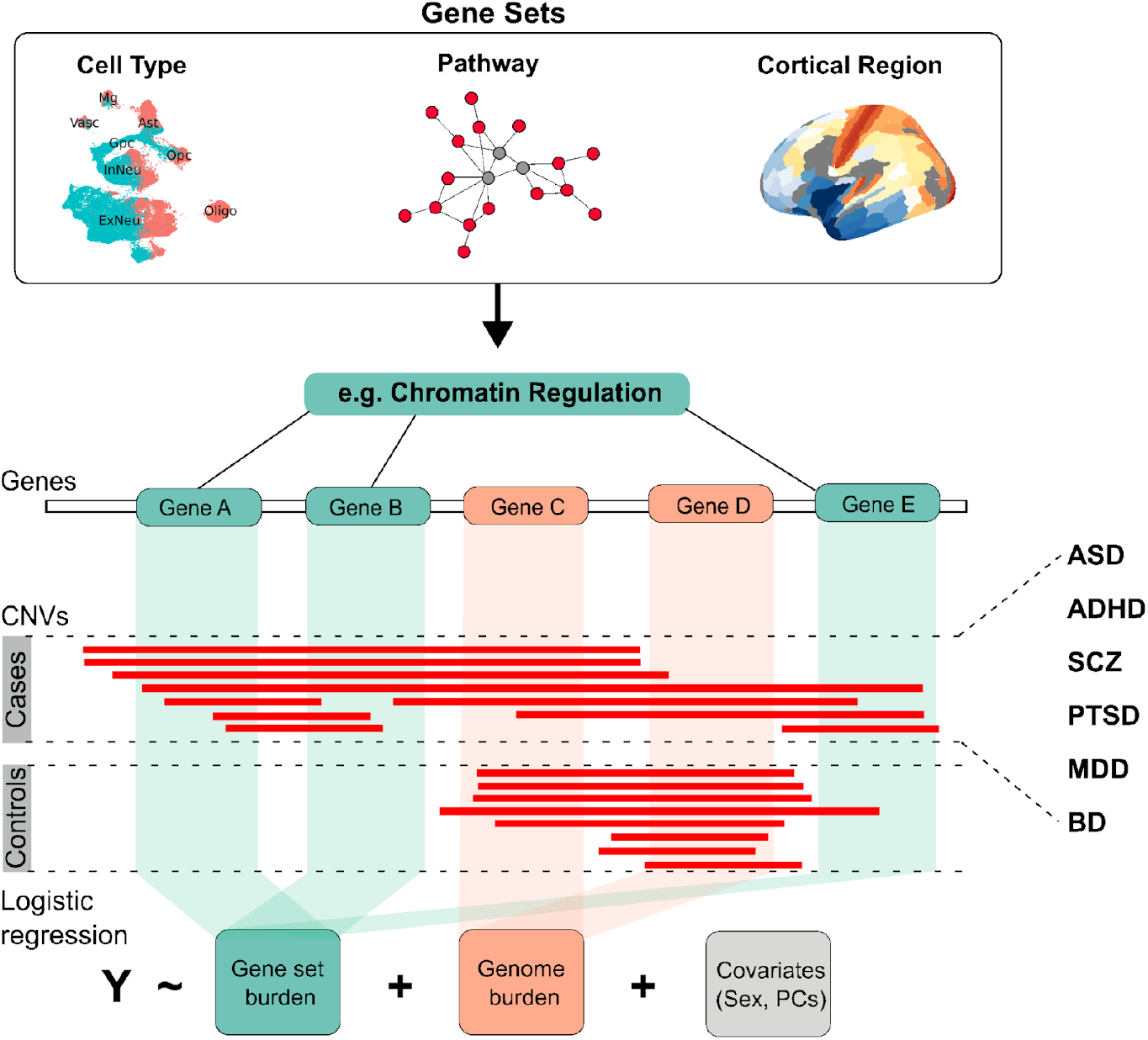
Investigating association of pathways, cell types and brain regions by Gene Set Burden Analysis (GSBA). Gene sets were derived for Pathway (from GO, KEGG, REACTOME, and BioCarta), Cell type (from single cell study, Velmeshev et al.), and Cortical regions (from Glasser parcellation of the Allen Brain Atlas). Case-control association of CNV burden collapsed across gene sets, was then tested by logistic regression and meta-analysis was performed across genotyping platforms. Functional gene set associations were tested for 6 major psychiatric conditions (ASD, ADHD, SCZ, PTSD, MDD, BD).

### Gene set association of rare CNVs in 6 psychiatric conditions

We leveraged large-scale rare CNV data (population frequency <2%) from the Psychiatric Genomics Consortium, comprising genome-wide microarray data from 574,965 individuals (133,007 cases and 441,958 controls) across six major psychiatric disorders: schizophrenia (SCZ), autism spectrum disorder (ASD), bipolar disorder (BD), major depressive disorder (MDD), post-traumatic stress disorder (PTSD), and attention-deficit/hyperactivity disorder (ADHD). CNVs were uniformly processed through a centralized pipeline for calling and quality control. Only rare CNVs (frequency <2%) were retained for analysis. Individuals represented diverse ancestral backgrounds, with 89.3% of European ancestry. This dataset enabled us to apply our multidimensional framework to identify distinct molecular and cellular features of brain function associated with each psychiatric diagnosis.

We assembled a primary catalogue of 2,645 gene sets that capture neurobiological features across multiple levels of organization. These included 2,453 molecular pathways from public databases ^41–43^. In addition, differential expression in single-cell expression data was analyzed to create gene sets for 12 cell types from human fetal and adult brain (ranging from second trimester to 54 years of age) ^44^, and differential expression in bulk tissue was analyzed to create 180 anatomic regions of cerebral cortex from the Allen Human Brain Atlas (AHBA) ^45^ ^46^(**Table S1**).

We investigated the association of functional gene sets with psychiatric diagnoses using **gene-set burden analysis (GSBA)** ^5^. Associations detected by GSBA capture the enrichment of variants in functionally-related genes in cases. However, GSBA is not equivalent to a gene-set enrichment test (e.g., Subramanian et al., 2005 ^47^). Rather, it is a statistical genetics approach that quantifies the effect size of rare-variant burden across a defined set of genes (e.g. a GO term) in cases and controls (**Fig. 1**). For each gene set, we tested the association of the aggregate DEL or DUP counts across genes with case-control status by logistic regression controlling for population structure, sex and overall genome-wide CNV burden (collapsed across all out-of-category genes). Gene-set summary statistics were generated for each genotyping platform in each diagnostic category, and results were combined by meta-analysis. Combined results were corrected for multiple testing with Benjamini-Hochberg False Discovery Rate (BH-FDR<5%, **Fig. S1**).

Significant functional burden associations were detected for a total of 787 gene sets in one or more conditions, including SCZ (671 gene sets) and ASD (331 gene sets), ADHD (52 gene sets), BD (122 gene sets) and MDD (3 gene sets) (**Table S2**). Comparing summary statistics between trans-ancestry analysis and the European-only subset, we found a high level of concordance in the z-statistics between single ancestry (European subset) and trans-ancestry results across the 6 psychiatric conditions (beta-coefficients are between 0.9 and 1 with median beta-coefficient = 0.97; **Fig. S2**). All results described below are from the trans-ancestry summary statistics, which has the greatest statistical power.

### Common neurodevelopmental pathways are implicated in multiple diagnostic categories

Pathway gene sets were compiled from Gene Ontology (GO) ^41^, KEGG ^42^, Reactome ^43^, and BioCarta ^48^, with size ranging from 50 to 500 genes. 589 gene sets were associated with one or more conditions (**Table S3**). Using Enrichment Map ^49^, overlapping gene sets implicated by CNVs were grouped into 19 functionally-related clusters representing canonical pathways such as MAPK signaling, nervous system development, synaptic transmission, chromatin regulation, etc. (**Fig. 2a, Table S4**). To summarize the pathway results, effect sizes were then estimated for the 19 gene sets in 6 diagnostic categories by GSBA regression (**Fig. 2b, Table S5**)

**Fig. 2:**
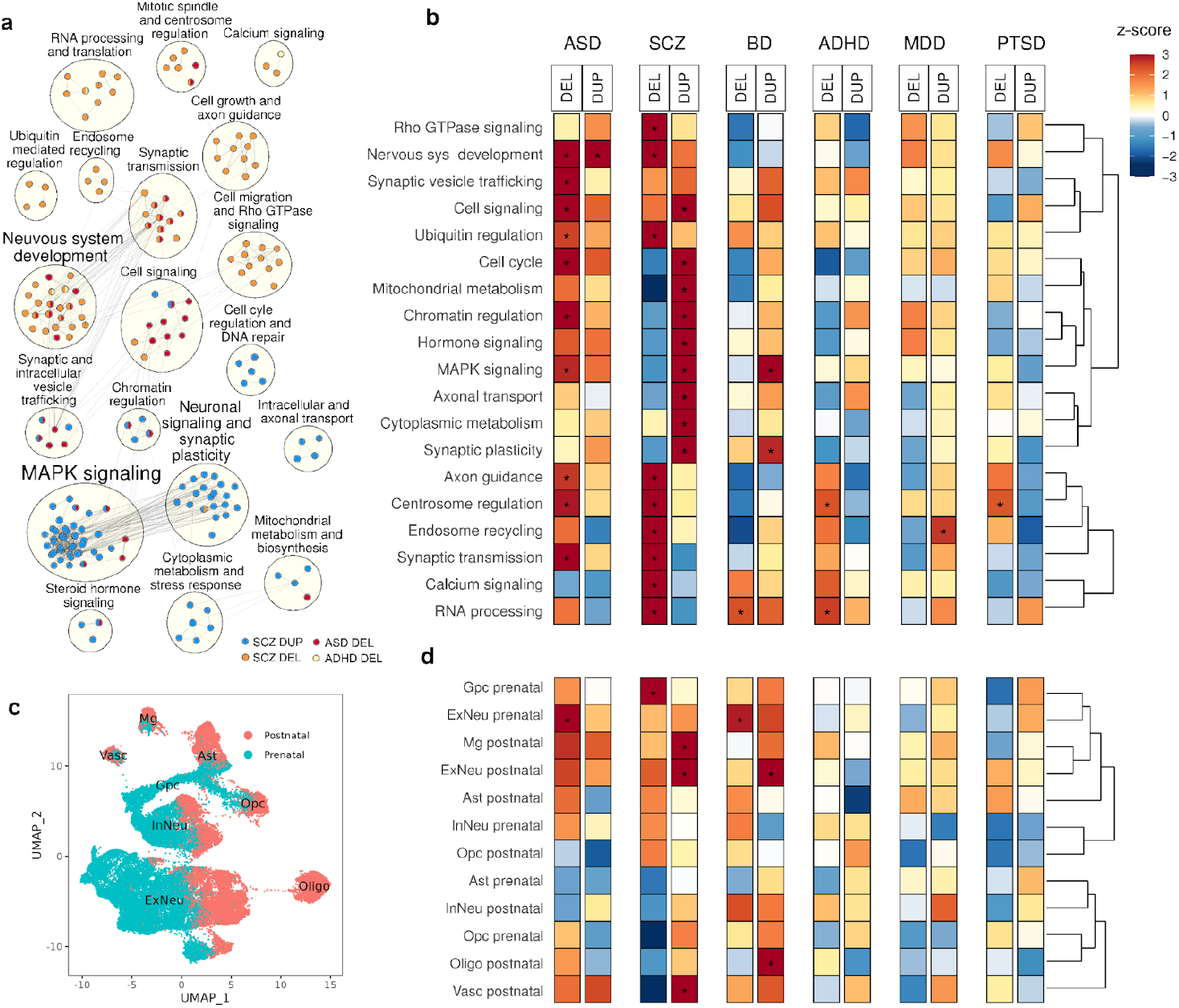
Rare CNVs association analysis results in molecular pathways and neuronal cell types. (**a**) Enrichment map showing clusters of functional modules that are significantly associated with any condition. CNV associations are color-coded as a portion with a node where red indicates a DEL association in ASD, orange indicates a DEL association in SCZ, blue indicates a DUP association in SCZ, and yellow indicates a DEL association in ADHD. Gene-sets not forming a cluster of 3 or more members were excluded. Gene set clusters are listed in **Table S4**. (**b**) The heatmap represents the results at the pathway-cluster level, with color indicating z-score from meta-analysis. (**c**) A UMAP plot displays cell clusters colored by prenatal (teal) and postnatal (red) periods. (**d**) Heatmaps show association results at the cell type level with color indicating z-score, where red represents a higher burden of CNVs in cases and blue represents a depletion of CNVs burden in cases. An asterisk indicates statistically significant associations (q-value <0.1). Summary statistics of the initial primary gene sets and for the final set of pathway clusters are in **Tables S3**, and **S5** respectively.

As expected, CNV burden associations were strongest in ASD and SCZ and were attenuated in other adult onset diagnoses, BD, ADHD, MDD, and PTSD. Many of the same functional gene sets were implicated in ASD and SCZ, including MAPK and other cell-signaling pathways, chromatin regulation, and synaptic transmission. Pathway signals in ASD were driven by significant DEL associations across 10 pathways. SCZ, by contrast, showed DUP associations in 9 functional gene sets such as chromatin regulation, MAPK signaling, axonal transport, and DEL associations in a different set of 9 pathways including synaptic transmission, axon guidance, and calcium signaling. The finding that pathway associations in SCZ differ by gene dosage is notable in light of the dose-dependent CNV effects reported for SCZ and other diagnoses in our companion study ^40^.

### Gene set burden associations implicate neuronal and non-neuronal cell types

Twelve cell-type gene sets were derived from single-cell RNA-sequencing of human cortex (prefrontal, cingulate, insula, motor, and temporal regions) spanning prenatal (5-9 months) and postnatal (0-54 years of age) developmental stages, based on the dataset from Velmeshev et al. ^44^. Starting from eight major cell type clusters defined in the original study, we refined these to capture key developmental distinctions, resulting in the following gene sets: five prenatal cell types - 1) glial progenitor cells (GpcPre), 2) oligodendrocyte precursor cells (OpcPre), 3) inhibitory neurons (InNeuPre), 4) excitatory neurons (ExNeuPre), and 5) astrocytes (AstPre); and seven postnatal cell types - 6) vascular cells (VascPost), 7) OpcPost, 8) oligodendrocytes (OligoPost), 9) microglia (MgPost), 10) inhibitory neurons (InNeuPost), 11) excitatory neurons (ExNeuPost), and 12) astrocytes (AstPost) (**Fig. 2c**). We observed several cell type associations with diagnostic categories (**Fig. 2d, Table S3**). ASD was associated with DEL burden in ExNeuPre, consistent with loss-of-function variants in ASD genes being enriched in fetal excitatory neurons ^7,50^. SCZ showed DUP association in ExNeuPre, microglia, and neurovascular cells and DEL association in GpcPre. BD showed DUP association in ExNeuPost and OligoPost and DEL association in ExNeuPre.

### Diagnoses differ in the distribution of gene-set associations between sensorimotor and association cortex

Spatial variation in gene expression across the cortex reflects region-specific regulation beyond differences in cell type composition ^51^. The primary gradient of gene expression (PC1) in the AHBA follows a sensorimotor-to-association (S-A) axis, spanning from primary sensory (visual, auditory, sensorimotor cortex) areas to transmodal (frontal, temporal) regions ^51–53^. This axis aligns with several cortical hierarchies, including developmental timing ^54–56^, myelination ^54,55^, anatomical projections ^57^, and functional specialization ^58^. Given its close correspondence with the S-A axis ^51^, we refer to AHBA PC1 as the S-A axis throughout the paper.

To investigate how gene dosage effects are distributed across the cortex, we defined gene sets for each of the 180 cortical regions from Glasser et al. ^45^. Gene expression was z-transformed across regions, and highly expressed genes (z>1) were assigned to each set of 180 regions. DEL and DUP burden was tested across cortical gene sets within each diagnosis. In total, 177 significant associations were identified. DEL and DUP associations are visualized on Glasser cortical maps (**Fig. 3a, c**), with effect sizes (z-scores) represented by a red-blue scale. We then tested whether spatial patterns of effect sizes aligned with the S-A axis using the SPIN test ^59^ with 10,000 permutations and Kendall correlation.

**Fig. 3:**
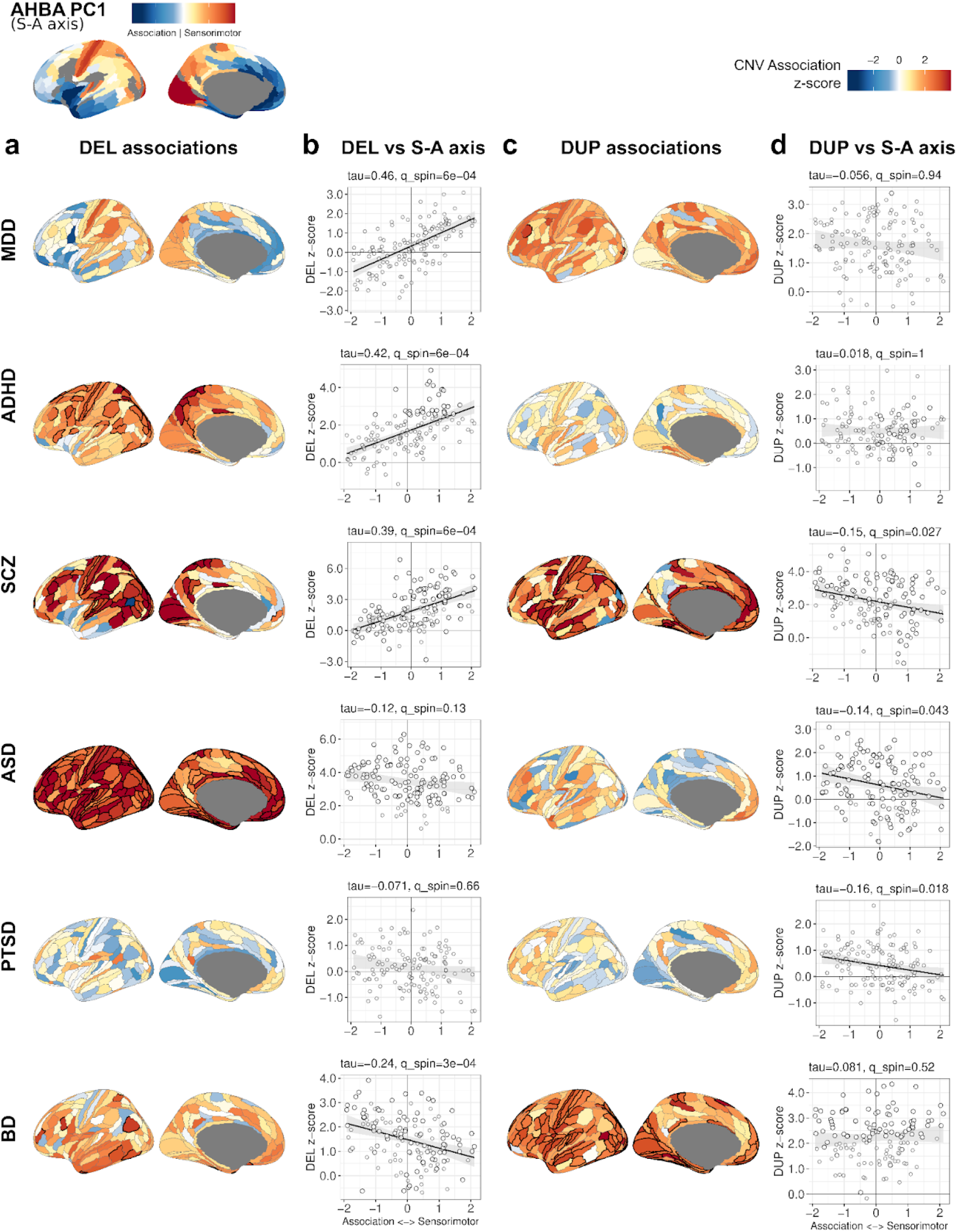
Rare (**a**) DEL and (**c**) DUP association analysis results of the cortical brain regions in the 6 conditions. Color indicates the association level (z-score) with red indicating the CNV association with the cases, while blue indicates the depletion of CNVs in cases (**Table S3**). Correlation results between CNV associations in (**b**) DEL and (**d**) DUP against the dominant transcriptomic brain gradient (PC1 of AHBA). Each circle represents a brain region gene set. Kendall’s Tau and corresponding q-value are shown in the title of each scatterplot. Solid diagonal trend line indicates significant correlation (q_SPIN_<0.05). The cortical map at the top left corner illustrates the transcriptomic gradient from PC1 AHBA.

CNV effect sizes varied across the cortex, and in several diagnostic categories, they showed significant, but divergent, correlations with the S-A axis. DEL effect sizes were positively correlated with the S-A axis in MDD, ADHD, and SCZ, indicating enrichment of DEL signal in sensorimotor cortex, while BD showed a negative correlation, indicating a relative enrichment of DEL signal in association cortex (**Fig. 3b**; **Table S3**). DUP associations were negatively correlated with the S-A axis in SCZ, ASD, and PTSD (**Fig. 3c,d**), indicating an enrichment in the association cortex. Our results suggest that the spatial distribution of gene dosage effects differs by diagnosis. Similar correlations were observed with other functional and anatomical gradients that are also aligned with the S-A axis (e.g., T1w/T2w ratio reflecting myelin content; **Fig. S3**) ^52^.

### Association of pathways with diagnosis varies by cell type and gene dosage

Our initial findings demonstrate that there are divergent genetic influences between different diagnostic categories when we stratify genetic effects by gene dosage and brain region. These findings highlight a principle that is somewhat obvious in retrospect. The multidimensional nature of psychopathology demands a multidimensional data analytic approach.

To characterize with more granularity how CNV effects are distributed in the brain, we investigated gene-dosage effects at the intersections of pathways, cell types, and brain regions. Pathway gene sets were intersected with cell types to create non-overlapping subsets (e.g., *Chromatin_ExNeu* and *Chromatin_InNeu*; **Fig. 4a**). Similarly, the transcriptome was divided into sensorimotor and association gene sets based on the correlations of individual genes with the S-A axis in the AHBA (76.29% of genes showed a nominally significant positive or negative correlation with PC1, **Tables S6-S7**). Pathways were intersected with these to create 2 region-specific subsets of each pathway (e.g., *Chromatin_Sensori, Chromatin_Assoc*). GSBA was then performed on two-way and three-way intersections of pathways (N = 19), cell types (N = 12), and brain regions (N = 2), including gene sets of size ≥30. Each gene set result was labeled with four factors: pathway, cell type, brain region, and dosage (**Table S8**).

**Fig. 4:**
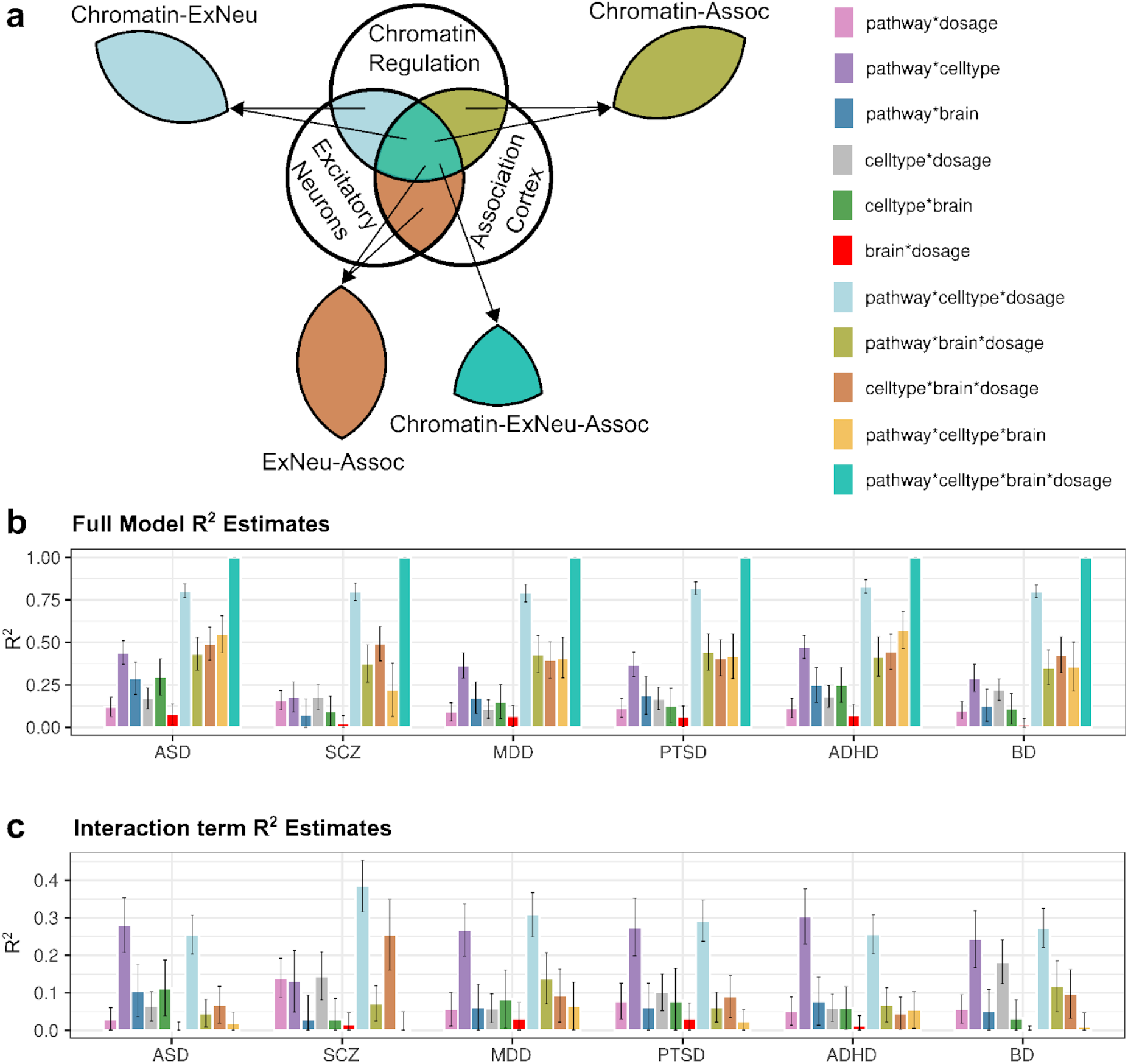
Associations of pathways with psychiatric traits vary by cell-type and gene dosage. (**a**) Schematic illustrating how gene sets were defined by intersecting pathway, cell type, and cortical region dimensions. Example intersections include Chromatin-ExNeu, Chromatin-Assoc, ExNeu-Assoc, and Chromatin-ExNeu-Assoc. (**b**) Full model R^2^ estimates showing the total variance in gene-set z-scores explained by main effects and interaction terms for each diagnosis. Models included pathway, cell type, brain region, dosage, and all combinations of two-way and three-way interactions. (**c**) R^2^ estimates for individual interaction terms, quantifying the contribution of each interaction to the explained variance. The pathway×celltype×dosage interaction consistently explains the largest proportion of variance across diagnoses, highlighting the importance of dosage-sensitive and cell-type-specific pathway effects (Tables S9-S10).

We then evaluated which levels of biological organization best explain variation in gene-set effects within each diagnosis. We performed linear modeling on effect sizes of stratified gene-sets (z-scores) with different combinations of pathway, cell type, brain, and dosage as independent variables. For each diagnostic category, variance explained (R^2^) in summary statistics was calculated for main effects and interactions of these factors. Of all 2-way combinations, pathway and cell type explained the greatest variance (35.3% on average across diagnoses, **Fig. 4b; Table S9**). A full model that further stratified gene sets by dosage explained a majority of the variance (80.1% on average). The pathway×celltype×dosage interaction consistently explained the largest proportion of variance (**Fig. 4c; Table S9**), explaining half of the effect of the full model. This result highlights the importance of cell-type-specific and dose-dependent pathway effects across psychiatric conditions. Model fits improved by 7-20% when the brain region was included in the models (**Fig. 4b,c; Tables S9-S10**), suggesting that spatial variation in pathway expression also explains a proportion of variance.

### Diagnostic categories are differentiated based on gene-dosage effects in pathways by cell type and brain region

To elucidate where gene-dosage effects converge at the intersection of pathways, cell types, brain regions, and psychiatric traits, we performed exploratory factor analysis (EFA) ^60^ of functional gene sets to identify latent factors that correspond to different gene-trait relationships. Genetic correlations of DEL and DUP associations across 6 diagnostic categories were estimated based on gene-set summary statistics (**Fig. 5a; Table S11**). Factor analysis of gene-set summary statistics was performed to extract latent dimensions of genetic effects, and a three-factor model was optimal (**Fig. S4**). Factor **F1** captured dose-dependent effects in SCZ and BD (DUP positive, DEL negative) and dose-aligned effects in ASD (DUP positive, DEL positive) in shared gene sets. **F2** captured DUP effects shared by mood disorders and PTSD. **F3** captured DEL effects shared by MDD, ADHD and SCZ. Importantly, genetic correlations between diagnostic categories show greater contrast when DEL and DUP results for each disorder were treated as independent components (**Table S11**) compared to when all gene set tests for DEL and DUP were aligned between disorders (**Fig. S5g; Table S12**). This result is consistent with diagnostic categories having involvement of common functional processes with sometimes opposing directionality. Loadings of DEL and DUP effects onto the 3 factors yields a unique profile for each diagnostic category (**Fig. 5b; Table S13**).

**Fig. 5:**
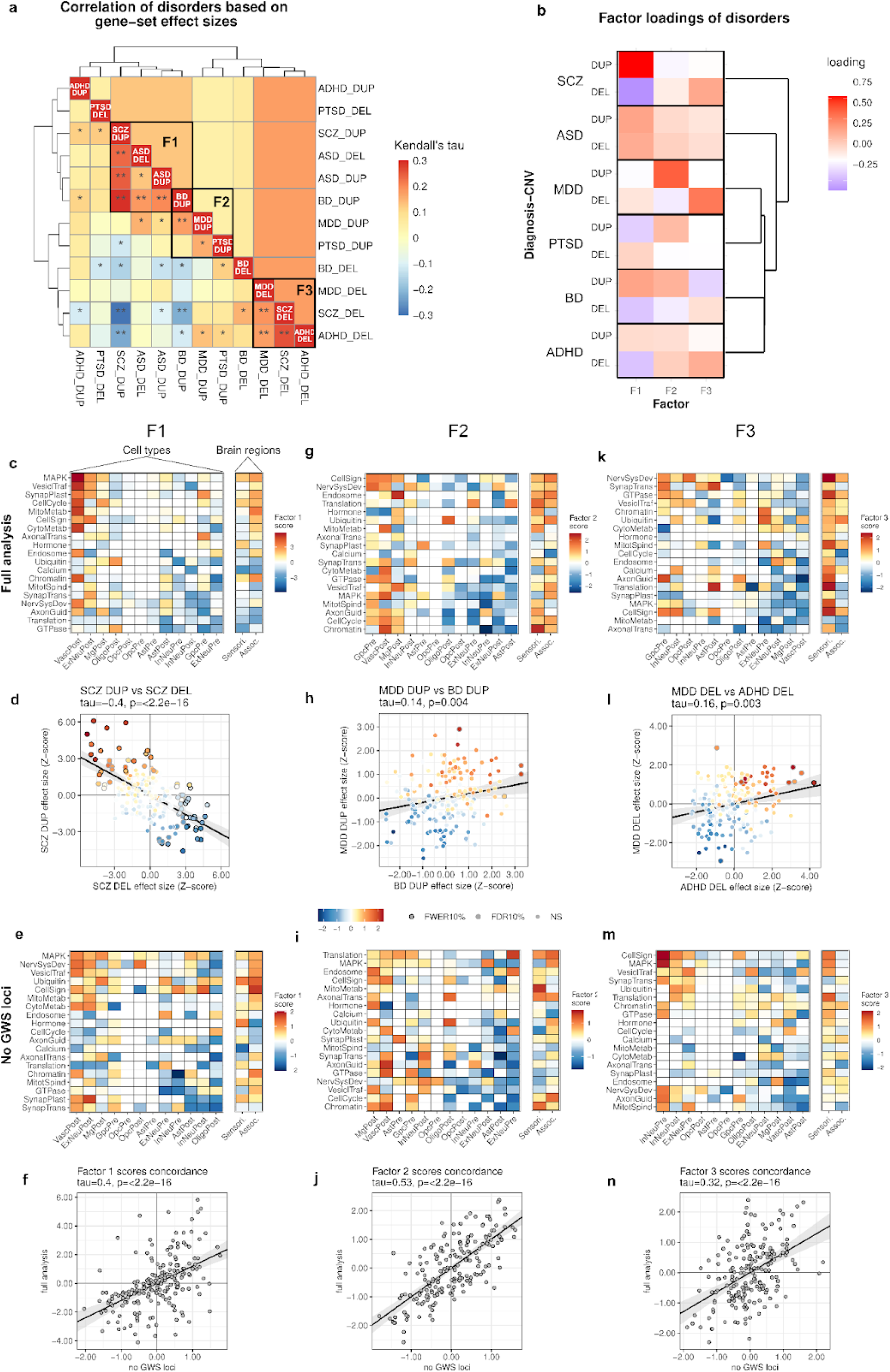
Differentiation of diagnostic categories based on gene-dosage effects in pathways by cell type and brain region. (**a**) Genetic correlations between diagnostic categories when each diagnosis-dosage combination is treated as an independent component, see also **Table S11**, *p<0.05) **q<0.05). Diagnosis-dosages with factor loadings >0.25 were grouped and labeled to highlight psychiatric traits contributing to F1, F2 and F3. (**b**): Factor loadings of DEL and DUP for disorders reveal a distinct profile for each diagnostic category. (**c, g, k**) Gene set-factor scores for the three factors, cell types and pathways were ordered using a simple sign-based bi-clustering algorithm (see methods) (Table S14). (**d**,**h**,**i**) Factor scores are representative of dose-dependent effects of genes. Scatterplots of gene set effect sizes (z-score) are shown for the top 2 diagnosis-dosage groupings with highest absolute factor loadings for factor F1, F2, and F3, and factor score of each gene set is indicated using the same color scale as in panels c,g,k. Solid trend lines indicate significant correlation between the diagnosis-dosage pair. (**e**,**j**,**m**) Factor analysis of gene sets with genome-wide significant loci removed yielded results with highly concordant gene set factor scores (**e**,**f**,**i**,**j**,**m**,**n**; tau_F1=0.45, tau_F2=0.53, tau_F3=0.32, p<2.2e-16; Table S14), demonstrating that these patterns are not attributable to a select subset of major loci.

The factor scores of functional gene sets show the relationships of neural processes to these latent dimensions. After a sign-based bi-clustering of the matrix, a structured pattern shows dose-dependent effects on pathways within cell types. **F1** in particular captures distinct clusters that represent the mirror-opposite effects of DUP and DEL seen in SCZ and other diagnostic categories (**Fig. 5b**). **Positively scoring gene sets** (**Fig. 5c**, upper left quadrant), which correspond to DUP associations in SCZ (**Fig. 5d**), were enriched for core regulatory processes (cell cycle, MAPK, chromatin) and metabolic pathways expressed in postnatal neurovascular cells (VascPost), excitatory neurons (ExNeuPost), and microglia (MgPost) (**Fig. S6**). **Negatively scoring gene sets** (**Fig. 5c**, lower right quadrant), which reflect DEL associations in SCZ (**Fig. 5d**), were enriched for calcium signaling, axon guidance, and translation pathways expressed in inhibitory neurons and glia. F1 Factor scores also reveal divergent effects on synaptic transmission by cell type, with DUP associations concentrated in excitatory neurons and DEL associations in inhibitory neurons, a pattern that is consistent with a shift in excitatory-inhibitory balance. To assess whether these patterns might be attributable to strong signals from a select subset of loci, we repeated GSBA (**Table S8**) and factor analysis (**Fig. 5e**) after removing 18 loci that reached genome-wide significance (GWS) in our companion study ^40^. The results showed highly concordant genetic correlations (**Fig. S5c**), factor solution and factor loadings (**Fig. S5f**), and gene-set factor scores (**Fig. 5f,i,l**). Thus the three factors derived in Figure 5 are not driven by a select subset of loci, and appear to be generalizable to CNVs genome wide. Similar clusters of pathway-cell type associations were evident in F1 (**Fig. 5e**), with the exception of the glial precursor cell type (GpcPre)(**Fig. 2d**). Lastly, F1 showed modest enrichment of gene set factor scores in Association cortex, a result that is consistent with the inverse dose-response of DEL (**Fig. 3b**)and DUP (**Fig. 3d**) effects along the S-A axis. Supplementary figures are provided that illustrate all gene-set associations (**Fig S6A**), the subsets that are captured by each of the latent factors (**Fig. S6B**), and functional terms that are enriched within each factor (**Fig. S6C**).

The orthogonal **F2** factor showed divergent positive (associated with cases) and negative (associated with controls) effects in developmental signaling (cell-cycle, MAPK, GTPase signaling) pathways in non-neuronal and neuronal cell types, respectively (**Fig. 5g,i**). **Positively-scoring gene sets**, which correspond to positive DUP associations in mood disorders (**Fig. 5h, Fig. S6b**), include nervous system development and metabolic pathways concentrated in microglia (MgPost), and neurovascular cells (VascPost). **Negatively-scoring gene sets** correspond to negative DUP associations in similar pathways in neuroectodermal lineages (ExNeuPre, ExNeuPost, InNeuPre, AstPost; **Fig. 5f,g**). The patterns in F2 suggest that DUP effects in mood disorders are concentrated in core regulatory processes in non-neuronal cell types, while DUP effects in core regulatory pathways may be tolerated (or protective) in neurons with respect to diagnoses of MDD and BD. Thus, DUP effects on regulatory pathways in postnatal excitatory neurons (e.g. GTPase_ExNeuPost, CellCycle_ExNeuPost, Chromatin_ExNeupost) are a point of divergence between F1 and F2 that represents neural processes that are positively associated with SCZ and ASD and not associated with mood disorders (**Fig. S6B-C**).

**F3** was characterized by positive loadings of MDD-DEL and ADHD-DEL (**Fig. 5b; Fig. S6b**). **Positively-scoring gene sets** consisted of DEL effects in Cell-signaling and neurotransmission (SynapTrans, VesiclTraff) in inhibitory neurons (InNeuPre, InNeuPost). **Negatively-scoring gene sets** were broadly distributed across regulatory and metabolic pathways in microglia and neurovascular cells. Notably, nearly all (18/19) canonical pathways showed strong positive F3 factor scores in the sensorimotor cortex (**Fig. 5i,k**), consistent with the positive correlation of MDD-DEL and ADHD-DEL with the S-A axis in **Figure 3a-b**. Thus, F3 captures differential DEL effects in synaptic and regulatory pathways that vary along the S-A axis and in cell-type populations that align with this cortical expression gradient, such as inhibitory interneurons ^55^.

### High-impact CNVs have a variety of cell-type specific gene-dosage effects

For CNV loci with the largest effect sizes on psychiatric traits, including reciprocal CNVs at 16p11.2, and 22q11.2 ^40^, clinical phenotypes are likely driven by the combined effects of multiple genes within each region ^39,61–63^. Results from this study further suggest that a CNV may exert its influence through distinct pathway effects in multiple cell types.

Duplication of 16p11.2 BP4-BP5 confers significant susceptibility to SCZ and BD, and Deletion is associated with ASD (**Fig. 6a**), consistent with some hallmarks of F1. Single-cell expression datasets ^44^ confirm that expression of genes within the locus differs significantly by cell type (**Fig. 6b**), A network was constructed representing cell-type expression of CNV genes and pathways (**Fig. 6c**), highlighting several pathway-cell type effects that are consistent with positively-scoring gene sets on factor F1 including several genes tied to regulatory pathways in neurovascular cells (MAPK3, ALDOA, MVP, TMEM219, TAOK2) and microglia (*CORO1A, INO80E*) as well as MAPK signaling and synaptic plasticity in postnatal excitatory neurons (YPEL3 PRRT2).

**Fig. 6:**
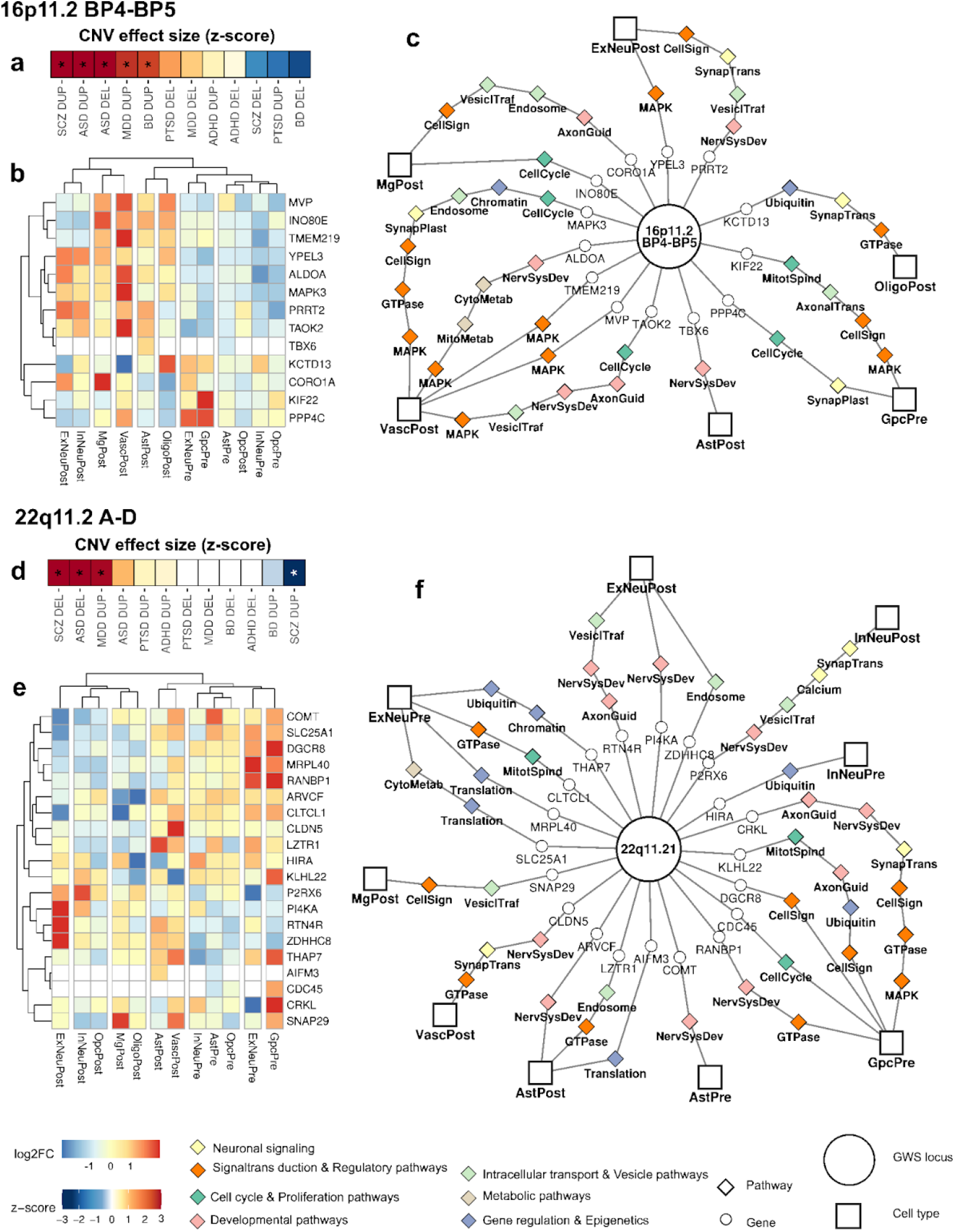
Cell-type specific expression of genes within major CNV loci 16p11.2 BP4-BP5 and 22q11.2 A-D suggests that the functional influence of a CNV in the brain may be driven by distinct pathway effects across a variety of cell types. CNV associations displayed in (**a**) and (**d**) were obtained from Shanta et al.^40^ Colors indicate the association direction and effect size (z-score), and asterisks indicate FDR<10% results. (**b**) and (**e**) heatmaps show log2 fold-change of cell type expression of the genes within each locus. The colors indicate the differential expression level. CNV-gene-gene-set networks in (**c**) and (**f**) display the CNV genes and their participation in the pathway-cell-type stratified gene sets. Shapes represent different entities of the network where the big circle in the middle is a GWS locus, peripheral circles are genes in the locus. A gene may be linked to one or more pathways (diamond) and at the end of the pathway, a cell type (square) is connected to indicate the gene membership of one or more stratified pathways of the same cell type. The color of diamond nodes indicates the group of pathways.

The 22q11.2 A-D locus has mirror positive and negative effects of DEL and DUP respectively on SCZ susceptibility (**Fig. 6d**), which is also a hallmark of F1. Pathway-cell type effects in 22q11.2 are consistent with negatively-scoring gene sets on F1, including chromatin, translation and GTPase signaling in fetal excitatory neurons (SLC25A1, MRPL40, CLTCL1, THAP7), axon guidance and endosome recycling in postnatal excitatory neurons (RTN4R, POI4KA, ZDHHC8) and calcium signaling in postnatal inhibitory neurons (P2RX6)(**Fig. 6e,f**). As mentioned previously, gene set effects listed here, persist after removing all genome-wide significant loci. Thus, the functional gene sets enriched within major CNV loci generalize to gene-dosage effects in the rest of the genome.

## Discussion

We present an integrative framework for characterizing the functional convergence and divergence of rare genetic influences on mental health traits. Using a statistical genetic approach, gene set burden analysis (GSBA) ^5^, we analyze the association of aggregate rare CNV burden in functional gene sets with diagnostic categories. A key element was to apply a multidimensional approach that quantified divergent effects of DEL and DUP in gene sets that represent the intersections of molecular pathways, neural cell types and cortical regions. This approach yields key insights into the neural basis of psychopathology. We demonstrate that, while major diagnostic categories converge on common molecular pathways, they diverge in the cellular context, spatial distribution, and directionality of genetic effects.

Gene-set burden tests identified 19 neurodevelopmental pathways, highly overlapping between ASD and SCZ, that were consistent with prior CNV ^3,5^, WES ^17,18^, and GWAS^15,64^ studies. These included pathways involved in neuronal signaling, GTPase and receptor mediated cell signaling, chromatin, translation, and metabolism. Cell-type associations included fetal excitatory neurons in ASD; excitatory neurons and oligodendrocytes in BD; and postnatal excitatory neurons, microglia, and neurovascular cells in SCZ. The involvement of neurovascular gene sets is notable given prior links of SCZ^65–67^, BD ^68^ and ASD ^32,69^ to cardiovascular disease. However, comparing lists of pathways and cell types does not reveal clear relationships between neural functions and diagnostic categories.

A key insight, originating from our companion paper ^40^, is the dose-dependent effect of genes in SCZ and other diagnostic categories, evident by the inverse correlation of effect sizes for reciprocal DEL and DUP of the same genes. Stratification of pathway associations by gene dosage showed that pathway associations, particularly in SCZ, differ by dosage. SCZ-DUP effects were concentrated in core regulatory pathways and DEL effects in neuronal signaling.

In addition, incorporating spatial patterns of gene expression into GSBA revealed differential genetic effects across brain regions. In several diagnostic categories, the spatial distribution of gene dosage effects aligned with the S-A axis, a cortical gene expression gradient, extending from transmodal association areas (frontal, temporal cortex) to sensorimotor regions (visual, auditory cortex), and spatial distributions differed by diagnostic category, with DEL effects in MDD, ADHD and SCZ enriched in sensorimotor cortex, while DEL effects BD and DUP effects in ASD, PTSD and SCZ were enriched in association cortex.

These findings highlight how stratification of genetic effects by context and gene dosage allow for the differentiation of diagnostic categories. To determine where genetic effects converge and diverge at multiple levels, we investigated gene-dosage effects in the interactions of pathways, cell types and cortical regions. Mixed-effects modeling demonstrated that associations of gene sets captured the largest share of variance when pathways were stratified by cell type, and dosage. Spatial information also contributed a modest additive effect representing differential genetic effects along the S-A axis, as observed for MDD-DEL and ADHD-DEL (**Fig. 3**).

Factor analysis revealed three latent dimensions of gene-dosage effects (F1, F2, F3) that capture shared and distinct genetic architectures across diagnoses. A major factor **F1** captured a set of neural processes that have a dose-dependent relationship to SCZ (DUP positive, DEL negative) and dose-aligned relationship to ASD (DUP positive, DEL positive), with *distinct pathway-cell type combinations at opposing ends of the dose-response curve*. SCZ-DUP associations in cell-signaling (MAPK, cell-cycle) and metabolic pathways were concentrated in postnatal excitatory neurons and neurovascular cells. SCZ-DEL associations in neuronal signaling (synaptic, calcium) were concentrated in inhibitory interneurons, consistent with an imbalance of excitation and inhibition ^70^. Dose-dependent effects in SCZ also correlated with the S-A axis (Fig. 3) with DUP effects aligned to the association cortex and DEL effects in sensorimotor regions. This pattern suggests that one major dimension of psychosis consists of negative effects on inhibitory activity (disinhibition) in sensory processing and positive dysregulation of excitatory processes in frontal/temporal regions. Thus, our genetic findings could inform studies of neurophysiology in schizophrenia ^71,72^. Notably, ASD contrasts with SCZ in the directionality of effects in F1. In contrast to the dose-dependent effects in SCZ, In ASD, opposing effects of DUP and DEL are concentrated within the same neural processes. This fact could reflect distinct linear and non-linear dose responses for the cognitive traits underlying psychosis and social behavior respectively.

Additional factors captured orthogonal neural processes associated with mood disorders and ADHD. **F2** implicated cell-type specific effects in mood disorders consisting of divergent positive and negative effects on cell-signaling between non-neuronal and neuronal cells respectively, the latter being a point of divergence from SCZ and ASD. These findings represent a possible genetic basis for differences in the densities of neurons and glia that have been reported in postmortem studies of BD and MDD ^26^. **F3** reflected differential DEL effects along the S-A axis capturing broad sensorimotor enrichment in ADHD and MDD consisting of synaptic and regulatory pathways in cell-type populations that align with this cortical gene expression gradient, such as inhibitory interneurons ^55^.

We also show that specific high-impact CNVs are enriched for combinations of cell-type-specific genes involved in pathways consistent with our broader findings. 16p11.2 BP4-BP5 ^4^ represents a genomic region that is enriched for multiple functional gene sets at the positive end of factor F1 (cell signaling pathways in ExNeuPost and VascPost). Conversely 22q11.2 A-D ^73^ is enriched for functional gene sets at the negative end of F1, such as regulatory pathways in ExNeuPre and calcium signaling InNeuPost. These results suggests that the large effects of an individual CNV may result from the combined impact of genes acting across multiple neural processes. Thus, 16p11.2 and 22q11.2 CNVs are monogenetic conditions that could serve as models for the dose-dependent effects of the major factor F1. High-risk CNVs, such as these represent patient groups that can be recruited for deep phenotypic characterization ^74^ and parallel functional characterization of neural processes in brain organoid models ^75,76^. Thus the findings from this study can be directly applied in clinical and translational studies of CNVs.

Our results provide a genetic basis for previous findings from other NIH-funded collaborations such as the PsychEncode consortium. Consistent with findings from Gandal et al., functional analysis of CNVs shows that core molecular pathways are shared by multiple diagnostic categories, such as ASD, SCZ, BD and MDD including synaptic transmission and neuronal signaling pathways ^19^ and there are divergent effects in neuronal and non-neuronal cell types ^20^. Considering just one level of biological organization at a time, such as pathways, the patterns that emerge from PsychEncode, GWAS, WES and CNVs are dominated broadly by “functional convergence” that seemingly spans all diagnostic boundaries. However, when genomic approaches take into consideration the joint influences of cell types, spatial distribution and directionality (dosage) of the pathway effects, distinct mechanisms emerge that underlie different dimensions of psychopathology.

## Data Availability

All data produced in the present study are available upon reasonable request to the authors

## Acknowledgements

We thank the many research participants. The PGC is supported by grants to UCSD (R01MH124847), UNC (R01MH124871), MGH (R01MH124851), Mount Sinai School of Medicine (R01MH124839), Cardiff University (R01MH124873), Trinity College Dublin (R01MH124875) and Washington University St. Louis (R01DA054869). Additional support was provided by grants to J.S. (MH119746, MH133899), C.N. (MH106595, MH124847) and S.J. (U01 MH119690, U01 MH119739, CIHR_495906). Funding for the work in Bipolar Disorder was supported by the Research Council of Norway (#223273, 248778, 262656, 273291, 283798, 248828), South East Norway Health Authority (2017-112), and KG Jebsen Stiftelsen. The iPSYCH project is supported by grants from the Lundbeck Foundation (R165-2013-15320, R102-A9118, R155-2014-1724 and R248-2017-2003) and the universities and university hospitals of Aarhus and Copenhagen. Genotyping of iPSYCH samples was supported by grants from the Lundbeck Foundation, the Stanley Foundation, the Simons Foundation (SFARI 311789 to M.J.D.), and NIMH (5U01MH094432-02 to M.J.D.). The Danish National Biobank resource was supported by the Novo Nordisk Foundation. Data handling and analysis on the GenomeDK HPC facility was supported by NIMH (1U01MH109514-01 to A.D.B.). High-performance computer capacity for handling and statistical analysis of iPSYCH data on the GenomeDK HPC facility was provided by the Center for Genomics and Personalized Medicine and the Centre for Integrative Sequencing, iSEQ, Aarhus University, Denmark (grant to ADB). Additional support to S.W.S from The University of Toronto McLaughlin Centre, the Hospital for Sick Children (SickKids) Foundation, the Ontario Brain Institute, Genome Canada/Ontario Genomics Institute and the Northbridge Chair in Paediatric Research held at the Hospital for Sick Children and University of Toronto.

## Competing interests

S.W.S. has served on the Scientific Advisory Committee of Population Bio and has been involved in Deep Genomics. Intellectual property from aspects of his research held at the Hospital for Sick Children are licensed to Athena Diagnostics and Population Bio.

## Author Contributions

The multidimensional analysis framework described here was developed through a team effort. Data analysis and manuscript preparation was led by WE and JS. Statistical models for multidimensional analysis of gene dosage effects were developed by WE and JS. Study design and statistical methods for meta-analysis of CNV across diagnostic categories was developed by OS and JS. Methods for spatial mapping of gene set associations were developed by KK and SJ ^77^. Statistical models for pathway analysis of CNV burden were developed DM, WE and SWS ^5^. CNV calling and derivation of CNV QC metrics and SNP-based ancestry PCs was performed by OS, BT, JM. Management of data access and enormous amounts of data wrangling were led by OH and OS in coordination with the remaining coauthors who carried out data collection.

## Data and code Availability

A WDL workflow containing all steps of CNV calling, QC and CNV-GWAS and meta-analysis code is under construction and will be released on the PGC CNV Github in conjunction with this publication (https://github.com/orgs/psychiatric-genomics-consortium/teams/cnv). Analysis code for GSBA and downstream analyses (https://github.com/naibank/PGC_GSBA) Gene sets, see Table S1, Gene-set summary statistics, see Table S3. Raw genotype and intensity files are available on subset of the cohort PGC dbGAP datasets https://www.ncbi.nlm.nih.gov/projects/gap/cgi-bin/collection.cgi?study_id=phs001254.v1.p1

Simons Foundation Autism Research Initiative SFARI (SSC and SPARK) https://base.sfari.org/

## Methods

### 1. Participants and CNV data

The CNV subgroup of the Psychiatric Genomics Consortium (PGC) works in collaboration with principal investigators from many labs to obtain large sample sizes of microarray data and analyze them using a centralized pipeline. We acquired microarray intensity files from GWAS for a total of 574,965 samples that included data from cases and controls for 6 diagnostic categories (Table S1 in our companion paper^40^). These samples were genotyped on 25 platforms across 4 genome builds. Data from Illumina was collected as either raw intensity data (IDAT) files or final report files while data from Affymetrix was collected as CEL files. To harmonize data, probes for newly acquired datasets were lifted over to GRCH38 for CNV calling while previously called CNVs were lifted over to GRCH38. Samples were genotyped on either Illumina or Affymetrix array.

For samples that were provided as IDAT files, the Illumina command line version of Genome Studio was used in conjunction with platform-specific manifest and cluster files to produce genotype call (GTC) files. Relevant features were extracted from GTC files to obtain final report files with probes, genotypes, Log R Ratio (LRR), and B Allele Frequency (BAF) for each sample. For samples that were not mapped to GRCH38, probe genome positions were converted to hg38 using the LiftOver tool. Samples within each platform were grouped into batches by plate. For Illumina/PsychChip arrays, CNVs were called using two methods: PennCNV and iPattern. For Affy6 arrays, CNVs were called using four methods: PennCNV, iPattern, CScore, and Birdsuite. For Affy5 and Affy500K arrays, CNVs were called using two methods: PennCNV and Birdsuite. For Axiom arrays, CNVs were called using two methods: PennCNV and QuantiSNP. The consensus of CNV calls from multiple callers was created by merging CNVs at the sample level and retaining CNVs that were called by at least 2 methods.

#### 1.1 Sample QC

Quality control (QC) was performed first at the sample level, and conducted independently for each microarray platform,according to methods from our previous CNV GWAS of schizophrenia (Marshall et al. 2017^5^). For Illumina arrays, LRR standard deviation, BAF standard deviation, and GC waviness factor were extracted from PennCNV log files. Samples were retained if each of the measures were within 3 SD of the median. Affymetrix arrays used MAPD and waviness-sd parameters from affy power tools. Samples were further evaluated based on the number and total length of autosomal CNVs detected, and were retained if these values did not exceed 3 SD of the mean. The proportion of the chromosome that was tagged as a CNV was calculated and samples were excluded if >10% of the chromosome was marked as a CNV region to filter possible aneuploidies.

#### 1.2 CNV QC

Large CNVs that were fragmented were merged. CNVs <10kb in length or containing <10 probes were excluded. CNV calls were removed if they spanned the centromere or telomere (100kb from end of chromosome) or had >50% overlap with segmental duplications, immunoglobulin, or T cell receptor (recurrent CNVs were processed without segmental duplications, immunoglobulin, and T cell receptor filters). The call set was restricted to rare CNVs with ≤10% frequency within-platform or across all platforms.

### 2. Ancestry Principal Components and Ancestry Partitioning

We extracted a subset of SNPs with < 1% missingness across all platforms (12,185 SNPs) and performed a principal component analysis using the flashPCA software^78^. In order to genetically infer the ancestry of each individual, we used the SNPweights software ^79^ on the same subset of SNPs to calculate % ancestry based on a reference panel containing 6 different populations (751 EUR, 687 EAS, 630 SAS, 568 AFR, 41 AMR, 22 OCE). Samples were categorized into 5 large homogeneous groupings based on the following criteria used in a previous study ^80^ 39 (Table S2, Fig S1): EUR: subjects with EUR ≥ 90%, AFR/AFAM: subjects with EUR < 90% & AFR ≥ 5% & EAS/SAS/AMR/OCE < 5%, ASN/ASAM: subjects with EUR < 90% & (EAS ≥ 5% or SAS ≥ 5%) & AFR/AMR/OCE < 5%, LAT: subjects with EUR < 90% & AMR ≥ 5% & EAS/SAS/AFR/OCE < 5% or EUR < 90% & AMR ≥ 60% & EAS < 20% & SAS < 15% & AFR/OCE < 5%, MIX: Uncategorized subjects.

### 3. Gene QC

To avoid having false positive findings arising due to a platform or dataset biases, we performed an extra filtering step of the genes being included in the gene set analysis. For each gene, separately for DELs and DUPs, CNV frequency was calculated per platform and dataset. Given the reduced penetration of the most recurrent CNVs, the incident frequency of such CNVs can be higher than that of disease prevalence. In particular, 15q11.2 DEL (major risk locus for ASD and SCZ) has been reported to have an incident rate between 0.57-1.27% ^81^, thus, using an inclusive frequency threshold, wWe then limited the CNVs to those with frequency lower than 2% across platforms and datasets. In addition, we calculated weight deviance score (WDS) of CNV frequency per platform/dataset and used that to derive a platform/dataset specificity index (SI). Specifically, for each gene, CNV frequency (C_i_) for a particular platform/dataset was compared to the expected CNV frequency (E_i_) estimated from across platforms/datasets as shown in Eq.1.

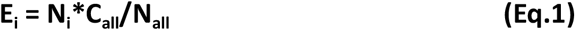

where for a particular platform/data i, E_i_ is the expected CNV frequency, N_i_ is the sample size, C_all_ is the CNV frequency in the entire dataset, and N_all_ is the entire dataset sample size.

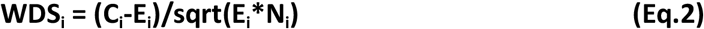

Then WDS_i_ was calculated as Eq. 2. With the max WDS across platforms/datasets representing the specificity index. We removed genes having dataset_SI ≥ 0.2 and platform_SI > 0.6 from subsequent analyses.

### 4. Gene set data

#### 4.1 Cortical regions

To generate gene sets for different cortical regions of the human brain, we acquired gene expression data in the brain from Allen Human Brain Atlas (AHBA; https://human.brain-map.org/static/download)^46^, multimodal brain parcellation from Glasser’s brain regions^45^. Using the Abagen toolbox (version 0.1.3; https://github.com/rmarkello/abagen)^82^, we mapped brain parcels and gene expression data, and then performed gene expression normalization and scaling. Specifically, a robust sigmoid function was used to normalize the expression data across genes to address inter-sample variation, while min-max normalization was applied after to scale the gene expression across tissue samples. Using the left hemisphere, we defined 180 regions from Glasser’s brain regions^83^. To generate the gene sets, the region-mean expression levels of each gene were z-transformed across the regions. Genes were then assigned to cortical region(s) when their z-score>1. The median gene set size was 4,429 genes (see **Table S1**). To visualize cortical region results, we used ggseg v1.6.5^84^ and ggsegGlasser R libraries for Glasser’s brain regions.

#### 4.2 Cell types

We obtained single-cell RNA-seq data from Velmeshev et al., 2023 ^44^, which contains the data >700,000 nuclei covering both prenatal and postnatal development periods and 8 defined cell type clusters. The 8 defined cell type clusters were 1. Oligodendrocyte precursor cells (OPC), 2. Vascular cells (Vasc), 3. Excitatory neurons (ExNeu), 4. Oligodendrocytes (Oligo), 5. Interneurons (InNeu), 6. Microglia (Mg), 7. Astrocytes (ASst), and 8. Glial progenitors (Gpc). Using the cluster result from the original study, we redefined the cluster by taking into account the developmental period of the cell. Doing so, we obtained 12 cell type clusters; 1. postnatal Opc, 2. postnatal Vasc, 3. postnatal ExNeu, 4. postnatal Oligo, 5. postnatal InNeu, 6. postnatal Mg, 7. postnatal Ast, 8. prenatal ExNeu, 9. prenatal Ast, 10. prenatal Opc, 11. prenatal InNeu, and 12. prenatal Gpc. We then generated cell type marker gene sets using FindAllMarkers() function from the Seurat package. Genes were assigned to a particular cell type cluster with the highest average log2 fold-change only when the corresponding p-value is < 0.05 (**Table S1**). The gene set size for cell types were smallest in prenatal OPC (181 genes), and largest in postnatal Mg (2,058 genes) with a median of 1,223 genes.

#### 4.3 Molecular pathways and pathway clusters defined using EnrichmentMap

We compiled gene sets from multiple databases including Gene Ontology ^41^, KEGG pathways^42^, and Reactome ^43^. We filtered the gene sets to include only those with size between 50 and 500 genes, excluding sets with broader definition (>500 genes) and those with low statistical power (<50 genes). In total, we acquired 2,453 gene sets. To reduce dependency between tests for multiple testing correction, we further exclude 758 more gene sets through a step-down approach. Specifically, for each gene set, we removed any smaller subset with substantial gene overlap (Jaccard’s index >0.75). The gene set sizes for molecular pathways range from 50 genes to 495 genes with a median of 145 genes.

To summarize the pathway associations, we applied the EnrichmentMap Cytoscape plugin ^49^ on the top associated gene sets (BH-FDR<5%, with z-score>0) from all the conditions. There were 361, 106, 7, and 5 gene sets associated with SCZ, ASD, BD, and ADHD, respectively. By limiting to pathway clusters with at least 3 gene set members, this results in 19 pathway clusters. We then constructed new gene sets by merging all gene sets within each cluster for subsequent analyses.

### 5. Gene set burden analysis and sample-weighted meta analysis

Differences in genotyping platforms have been known to confound CNV detection given the variance in probe coverage. While the most common way to tackle platform bias in CNV data analysis is to model the effect as one of the covariates, however, the effect is not well controlled in a single regression model. In this study, we performed gene set burden analysis independently for different genotyping platforms and meta-analyzed the summary statistics derived from the individual platform analysis. Using ASD and SCZ as a preliminary experiments, in both conditions, we found a smaller genomic inflation factor or lambda (λ) value (Eq.3) in the meta-analysis result (λ_ASD_=1.78, λ_SCZ_=3.35) compared to the mega-analysis result (using platform as a covariate, λ_ASD_=1.82, λ_SCZ_=3.66).

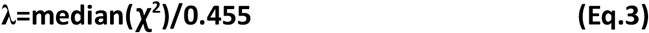

where χ^2^ is chi-square statistics, and 0.455 is the theoretical mean of chi-square distribution.

Specifically, we performed the gene set analysis on platforms where there are at least 50 cases and 50 controls. For each platform, a univariate analysis was conducted to compare the burden of genes in a gene set impacted by DELs or DUPs between cases and controls. The univariate analysis was done in one of two ways, either 1) a traditional case-control comparison for each individual condition, or 2) a family-based comparison. For the traditional case-control comparison, logistic regression was applied by regressing the number of genes in a gene set impacted by DELs or DUPs on the affection status (1 = affected, 0 = unaffected). Population structure (PC1-10), sex, and the number of genes outside the gene set impacted by DELs or DUPs were used as covariates to correct for any biases in the population, sex and total burden load. For the family-based comparison, we applied conditional logistic regression the same way logistic regression was applied, except that samples were matched by family ID. A likelihood ratio test was done to estimate p-value by comparing two regression models with and without the testing variable, in this case, a gene set burden.

A sample-weighted meta-analysis was applied to account for substantial differences in sample size between platforms. We derived the weight for each platform based on the effective sample size as shown in Eq.4.

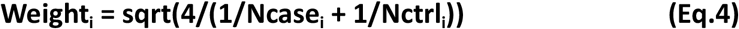

where Ncase_i_ is the number of cases in platform_i_, and Nctrl_i_ is the number of controls in platform_i_.

### 6. Gene burden analysis

We generated gene-level summary statistics by meta-analyzing the summary statistics from individual platform gene burden analysis. Similar to the gene set burden analysis, the gene burden analysis was done by either performing a logistic regression for case-control dataset, or conditional logistic regression for family-based dataset. We regressed the status of the CNV whether or not a sample has DELs or DUPs overlapping a particular gene on the affection status of the condition. Like gene set burden analysis, population structure (PC1-10), and sex were corrected in the analysis, with family ID being a random effect variable for conditional logistic regression. As multigenic CNVs might drive correlation between tests and that would affect multiple testing correction, genes were merged when the Jaccard index estimated from the proportion of CNVs commonly found between genes was >0.75. Since we only used the gene burden results to visualize findings from the main analysis, we did not report them in this study.

### 7. Correlation analysis of CNV association and Sensorimotor-Association axis and pathway-S-A-axis gene set stratification

We investigated how CNV associations distributed along the cortical gradient using the dominant brain transcriptomic variance data compiled in Dear et al ^51^. This is the PC1 of AHBA transcriptomic profile^46^ projected on the Glasser parcellation ^45^. The data was processed to exclude spatially inconsistent genes and, under sampling parcellations with a low number of donors (<6 donors). As a result, the final principal component analysis was performed on 134 parcellations and 7,937 genes. The CNV meta-analysis summary statistics of 134 Glasser parcellations was then compared with the PC1 AHBA using Spatial Permutation Inference (SPIN test^59^ with 10,000 permutations) with Kendall coefficient analysis.

To stratify gene set by the S-A axis, we first compute the Kendall coefficient of each gene against the PC1 AHBA. The gene expression matrix was preprocessed and obtained from Dear et al ^51^ where it contains the data for 10,028 genes, of which 8,588 genes are a member of at least one gene set. This identified ∼76% of the genes (n=6,552) to be correlated with the S-A axis at nominal significant level (p<0.05). We then stratified each gene set into 1) sensorimotor cortex set (tau>0, p<0.05), and 2) association cortex set (tau<0, p<0.05), leaving out other non-correlated genes from the subsequent analysis.

### 8. Genetic correlations based on gene-set summary statistics

We compared each pair of summary statistics (e.g., a pair of DEL and DUP summary statistics) 1) within the same condition to assess dosage sensitivity at the gene set level in each condition, and 2) between two conditions to assess gene set profile similarity between conditions. To do so, we performed a Kendall rank correlation analysis of the z-scores estimated from the meta-analysis of gene set burden results across individual platforms.

To examine correlations between cortical maps (e.g., CNV associations, transcriptomic gradient map, etc.), we applied a commonly used spatial Kendall’s correlation and assessed significance against a two-sided spatial autocorrelation-preserving null mode (SPIN test) ^59^, accounting for high inter-regional correlations as a result of spatial smoothing. To reduce the influence of gene set size on the z-score and the estimated correlation, we regressed out the gene set size from the z-score and performed correlation analysis on the residuals.

Using stratified gene set summary statistics, we estimated genetic correlations in 2 ways: (1) First, was to treat each diagnosis-dosage combination as an independent component and to examine the correlations of each (12 × 12, **Table S11**). (2) Second we examined the correlation of all the gene-set effects combined by stacking DEL and DUP summary statistics and aligning them directly between diagnoses (**Table S12**). Genetic correlations of independent diagnosis-dosage combinations showed greater contrast between diagnostic categories (**Fig. S5g**).

### 9. Latent factor analysis

We performed a latent factor analysis on the two-way (pathway-cell-type, pathway-brain) and three-way (pathway-cell-type-brain) stratified gene set summary statistics to investigate the shared and convergent dosage effects amongst the 6 psychiatric conditions using psych R library. The number of factors was optimized using a scree plot (elbow plot) of PCA on the summary statistic where a 3-factor solution was chosen (**Fig. S4**).

We specified a 3-factor solution using the fa() function, which estimates factor loadings that describe how each diagnosis-dosage combination contributes to the latent factors. A heatmap of the factor loading matrix, styled similarly to the genomic SEM plots, was generated to visualize how diagnoses align with these latent dimensions. To relate individual pathway gene sets to the latent factors, we computed the factor score for each gene set as a product between their z-scores and the factor loadings across diagnosis-dosage combinations, producing a second heatmap that highlights which biological pathways align most strongly with each latent genetic factor. For this second heatmap, specifically for pathway-cell-type stratified gene sets, we performed a sign-based bi-clustering on the heatmap where pathways are in rows, and cell types are in columns. The pathways and cell types were ordered in a descending order based on their average factor scores. This resulted in two main clusters of groups of pathways and cell types for i) positive and ii) negative factor scores.

All 2 way and 3 way stratification were included in the mixed-effects model analysis (**Fig. 4**). A factor analysis of three-way stratified gene sets (**Fig. S8)** produced similar factor solutions, genetic correlations and factor scores as the results in **Figure 5**. However, overall signal was comparatively weak due to the sparsity of the counts in the 3-way stratification of the data and the sparsity of gene sets that could be included in the analysis (stratifying pathway, by celltype, brain and dosage resulted in >50% of gene sets meeting the minimum size of 30 genes, **Fig. S7**). Thus main results in **Figure 5** include only the 2 way (pathway-cell type and pathway-brain) gene sets.

### 10. Linear model analysis investigating variance explained by different genetic factors

Using stratified gene set summary statistics, we evaluated which levels of biological organization best explain the variation in the gene-set effects within each diagnosis. We performed linear modeling on the effect sizes of the stratified gene-sets (z-scores) with different combinations of pathway, cell type, brain, and dosage as independent variables. For each diagnostic category, variance explained (*R*^*2*^) in summary statistics was calculated for the full model, the main effects, and the interactions of these factors. For example, suppose we would like to test for the effect of a cell type variable and its interaction term. Let *m0* be the full model of all three variables (e.g., *logit(y) ∼ pathway*celltype*dosage*), *m1* be the model without the interaction term with all three variables (e.g., *logit(y) ∼ pathway*celltype + celltype*dosage*), *m2* be the additive model without the evaluating variable (e.g., *logit(y) ∼ pathway + dosage*), and *m3* be the additive model with all three variables (e.g., *logit(y) ∼ pathway + celltype + dosage*). The *R*^*2*^ of the full model is from m0, the *R*^*2*^ of the main effect is estimated as *R*^*2*^*m3*-*R*^*2*^*m2*, and the *R*^*2*^ of the interaction term is estimated as *R*^*2*^*m0*-*R*^*2*^*m1*. A likelihood ratio test was performed to estimate the level of significance for each comparison through the *anova()* function in R.

## Supplementary materials

**Fig. S1:**
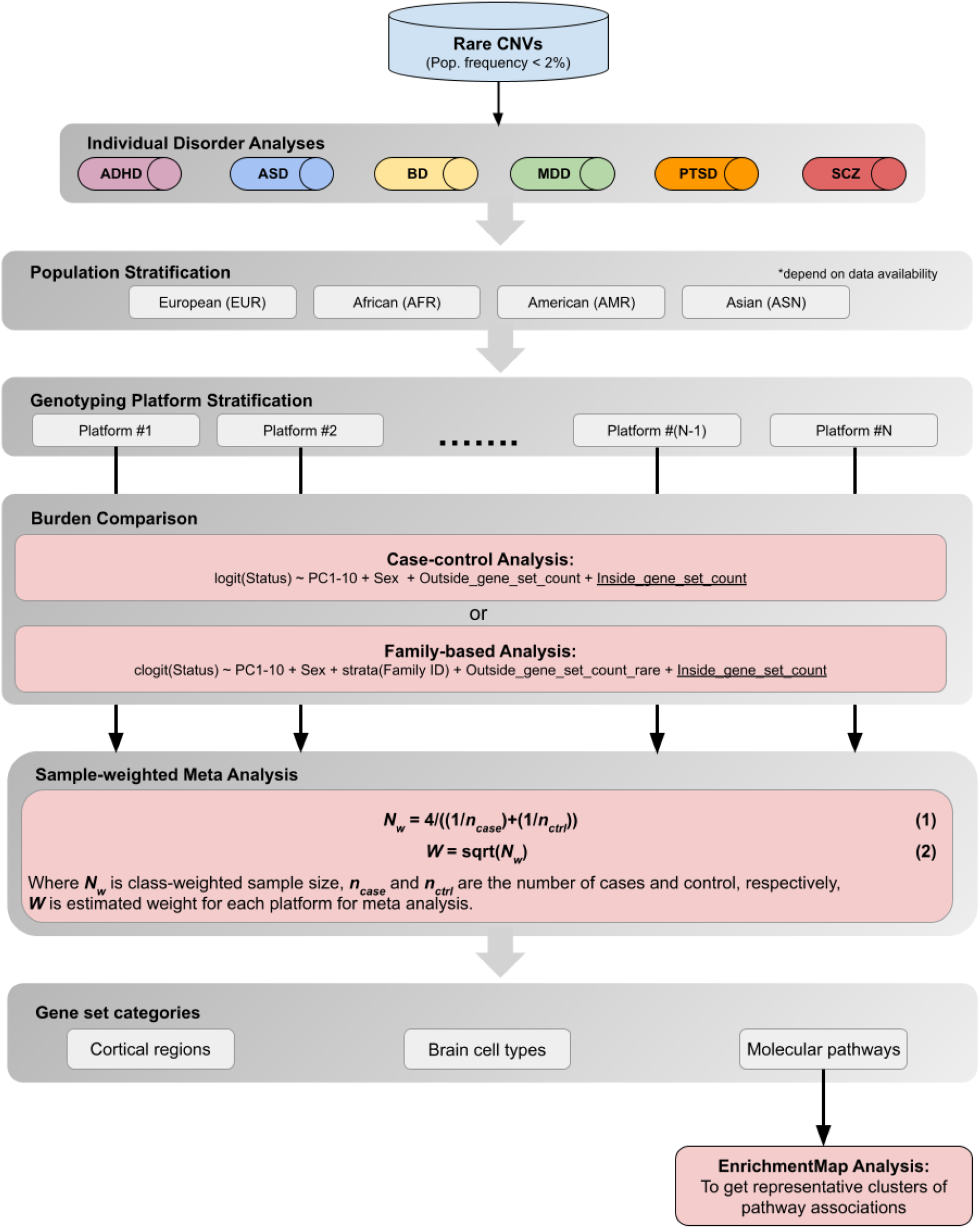
Gene set burden analysis (GSBA) workflow. A diagram showing the analytical procedure done for the gene set analysis of CNV data. First, CNVs were called and filtered down to rare CNVs at 2% frequency across platform and ancestry. Then, for each individual condition, to maximize the statistical power, we performed a cross-ancestry analysis, and also stratified the analysis by population groups; European (EUR), African (AFR), American (AMR), and Asian (ASN). For each stratified analysis, the gene-set burden comparison were done independently for each genotyping platform, then their summary statistics were meta-analyzed. For the burden comparison, we either performed a logistic regression for case-control data, or a conditional logistic regression for family-based data where family ID was used as a strata. Meta-analysis was done using a sample-weighted procedure (Eq.3-4), as it has shown a better robustness compared to a standard-error-based procedure. For the result of molecular pathways, we further clustered them using EnrichmentMap to obtain representative pathway clusters.

**Fig. S2:**
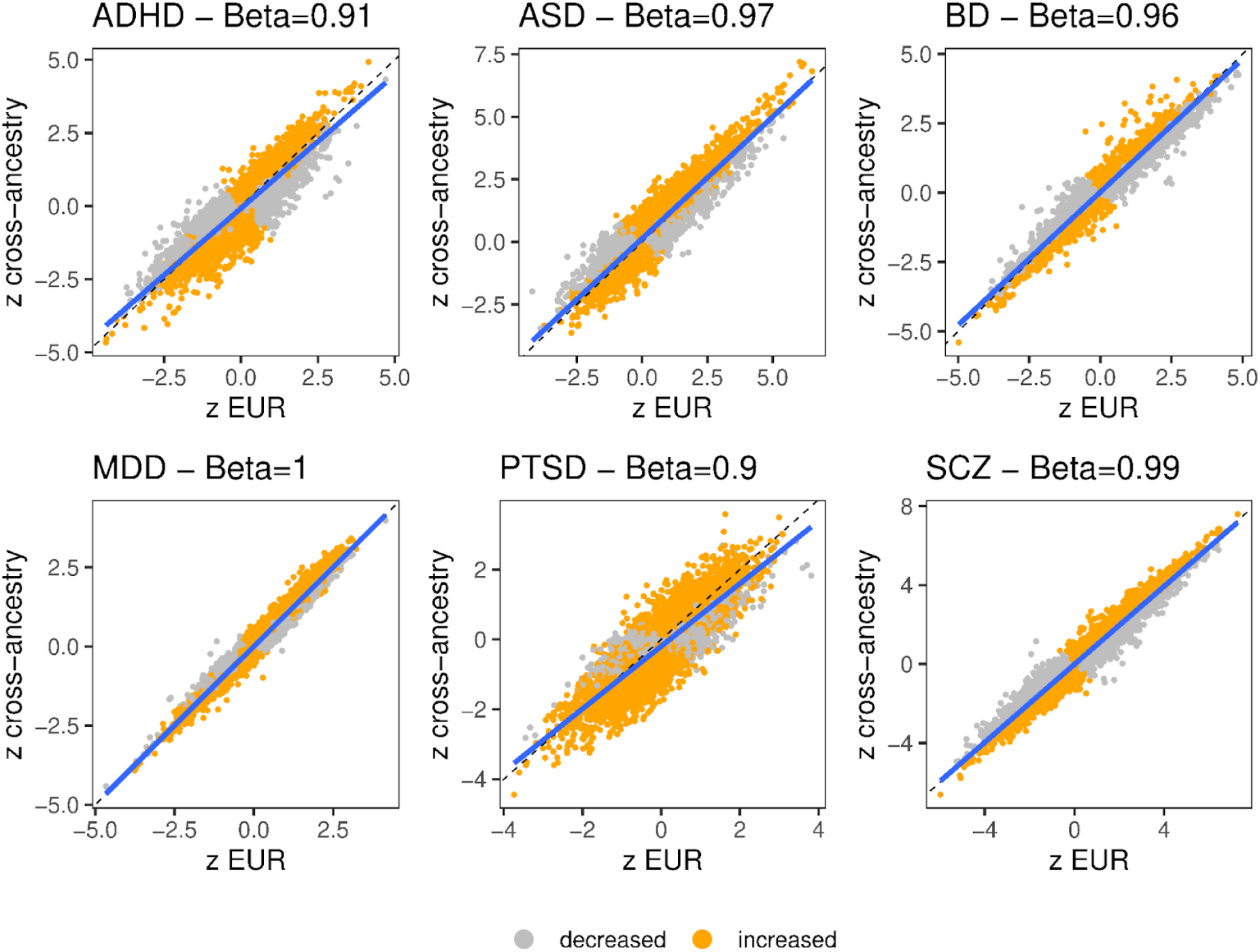
**Comparing GSBA results between the full cohort and subjects of only European ancestry** - scatter plots comparing summary statistics (z statistics from the sample-weighted meta analysis) between the analysis of European subset and the analysis of all ancestry. Beta coefficients estimated from linear model regressing z statistics from European analysis on the z statistics of cross-ancestry analysis.

**Fig. S3:**
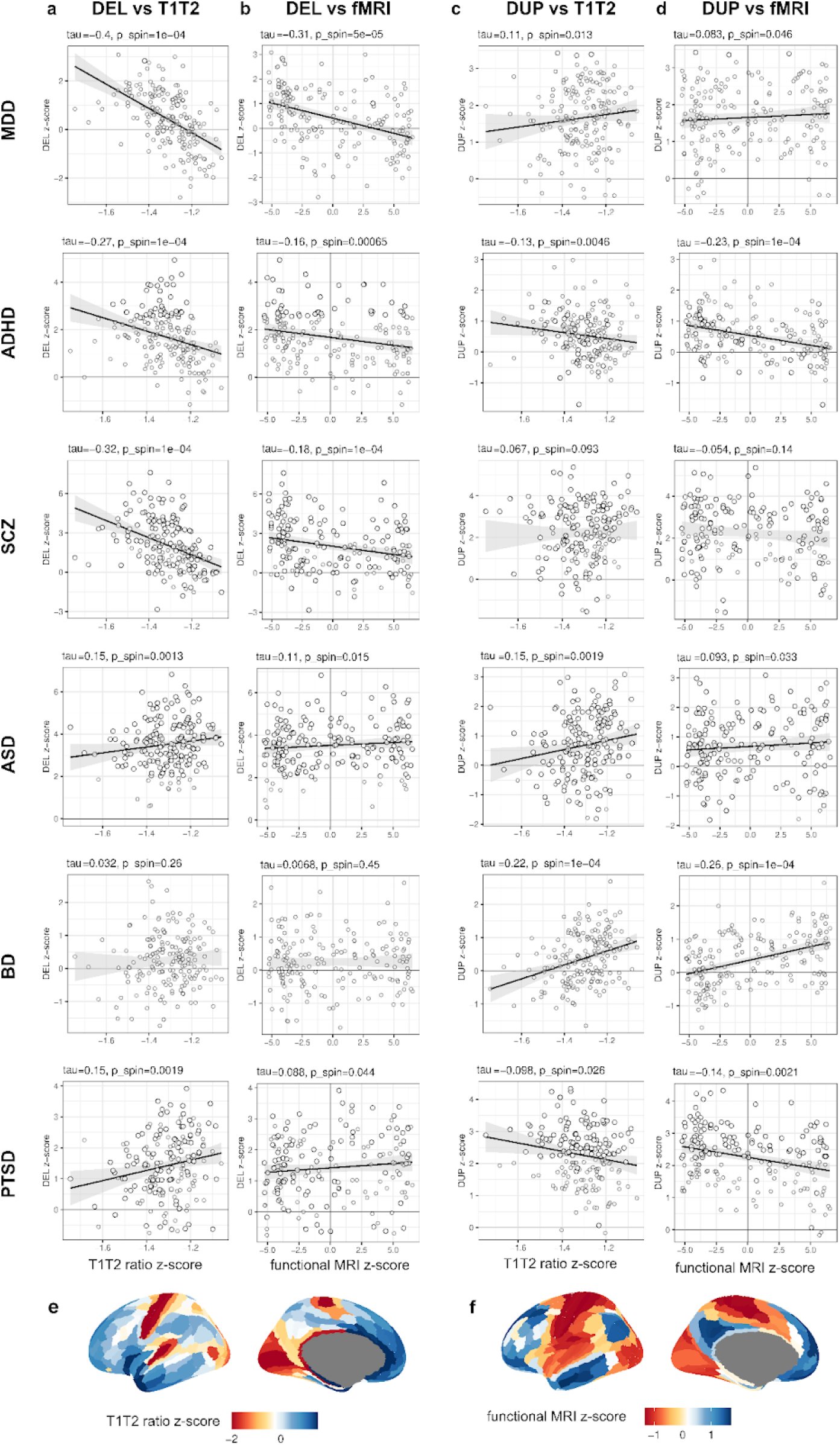
Correlation of cortical gene set effects with additional sensory–association cortical gradients. Scatter plots show the correlation between gene-set burden z-scores for DEL (**a**,**b**) and DUP (**c**,**d**) for two independent measures of cortical organization: the T1w/T2w ratio which reflects regional variation in intracortical myelination, and the principal gradient of resting-state fMRI. T1w/T2w and fMRI measures aligned to the Glasser grain maps were obtained from Markello et al. ^52^, both of which parallel the S-A axis derived from transcriptional principal components(Fig. 3). Correlation of CNV effects with these gradients supports the spatial specificity of gene-dosage associations across multiple cortical modalities. Together, these analyses highlight the convergent spatial patterning of CNV effects along major anatomical and functional cortical hierarchies. (**a**) DEL z-score and T1-T2 ratio, and (**b**) DEL z-score and fMRI. (**c**) DUP z-score and T1-T2 ratio, and (**d**) DUP z-score and fMRI. Solid trend lines indicate significant correlation where p_SPIN_<0.05. Brain maps of T1-T2 ratio and fMRI are shown in (**e**) and (**f**) where colors indicate the z-score.

**Fig. S4:**
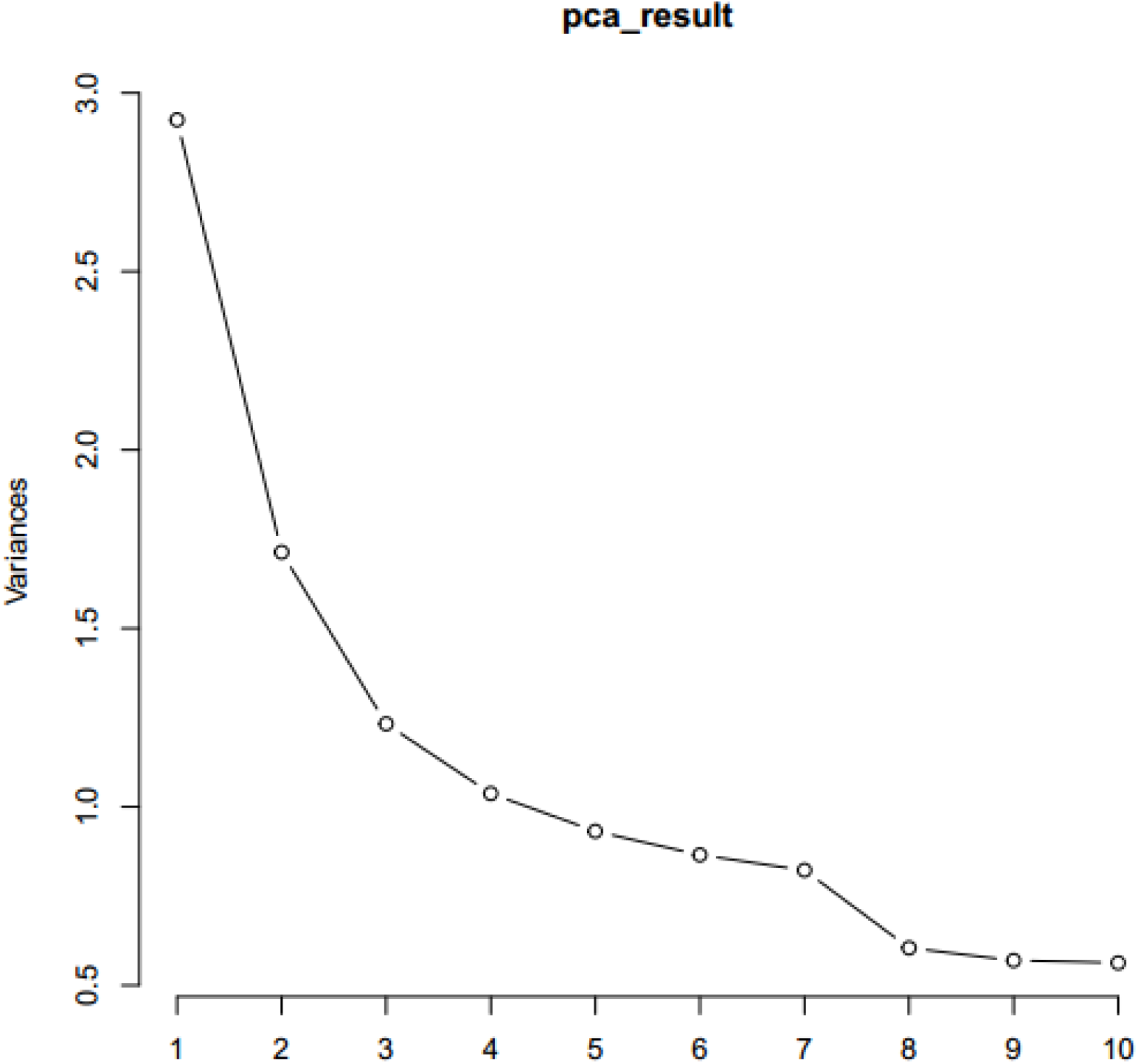
Using elbow plot (scree plot), we estimated an optimal number of factors to be 3 factors (variance drop threshold<5)

**Fig. S5:**
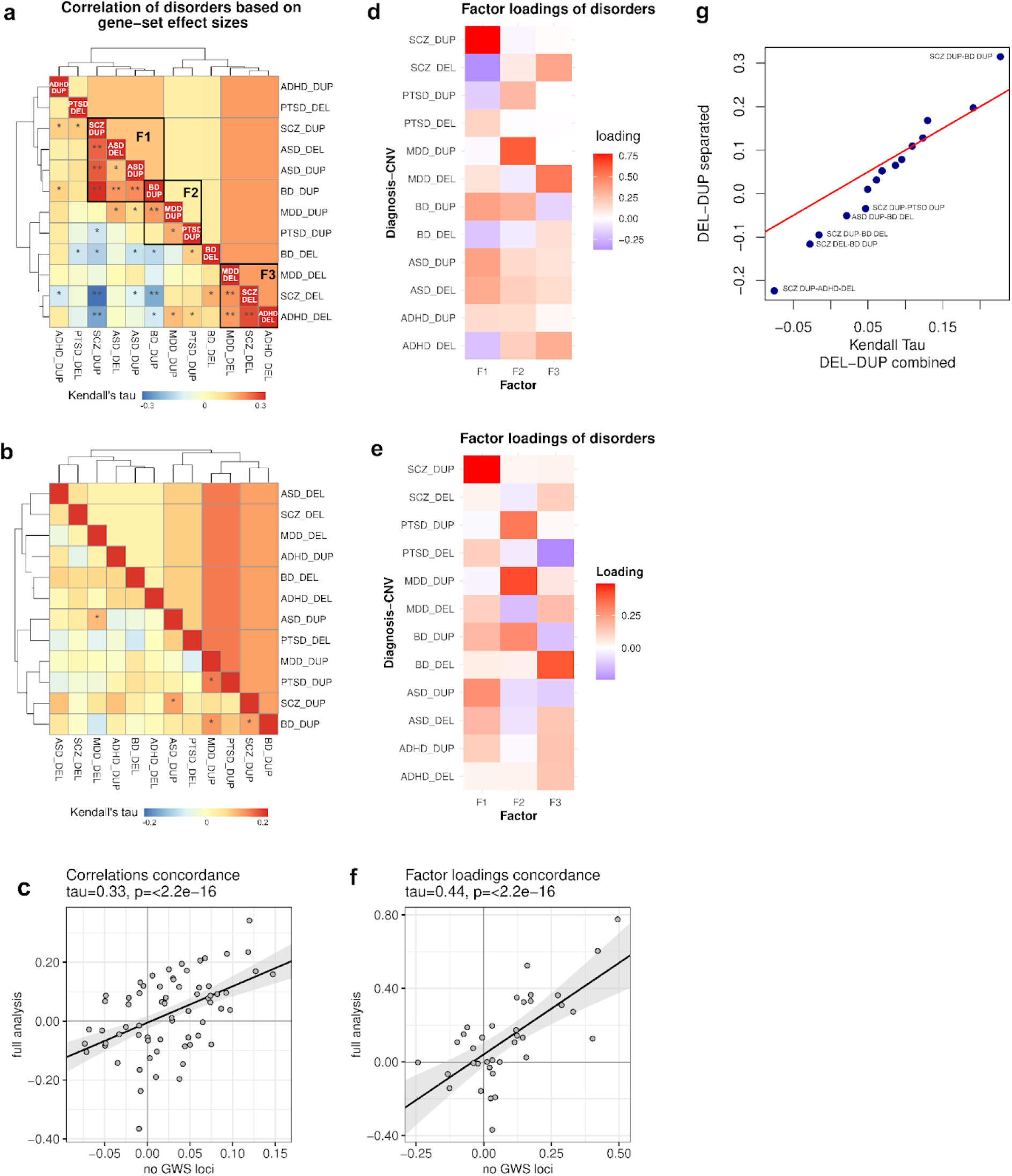
Factor analysis on two-way pathway stratification summary statistics without (by cell type, and by brain region) genome-wide significant (GWS) loci included in the analysis. Genetic correlations between diagnosis-dosage from (**a**) the full analysis and (**b**) the analysis without GWS loci. Single asterisks (*) indicate nominal significance (p<0.05), while double asterisks indicate significance after multiple testing correction (q<0.05), and a factor loading threshold of > 0.25 was applied to determine factor members. (**c**) Correlation of genetic correlation calculated from full analysis and no GWS loci analysis. Factor loadings of diagnoses reveal distinct signatures of diagnostic categories from (**d**) the full analysis and (**e**) no GWS loci analysis. (**f**) Correlation of factor loadings from full analysis and no GWS loci analysis. For (**c**) and (**f**) scatterplots, solid trend lines indicate significant correlation. Kendall’s Tau and corresponding p-value are reported in the title of the scatterplot. (**g**) QQ-plot comparing the distributions of correlation coefficients (Kendall’s Tau) when DEL and DUP effects in each diagnosis are treated as separate components (y-axis, Table S11) vs when the full sum stats of DEL and DUP are aligned between diagnoses (x-axis, Table S12). The negative tail of the y-axis distribution on the QQ plot was weakly skewed, suggesting that the distribution was enriched for effects that diverge between diagnoses.

**Fig. S6:**
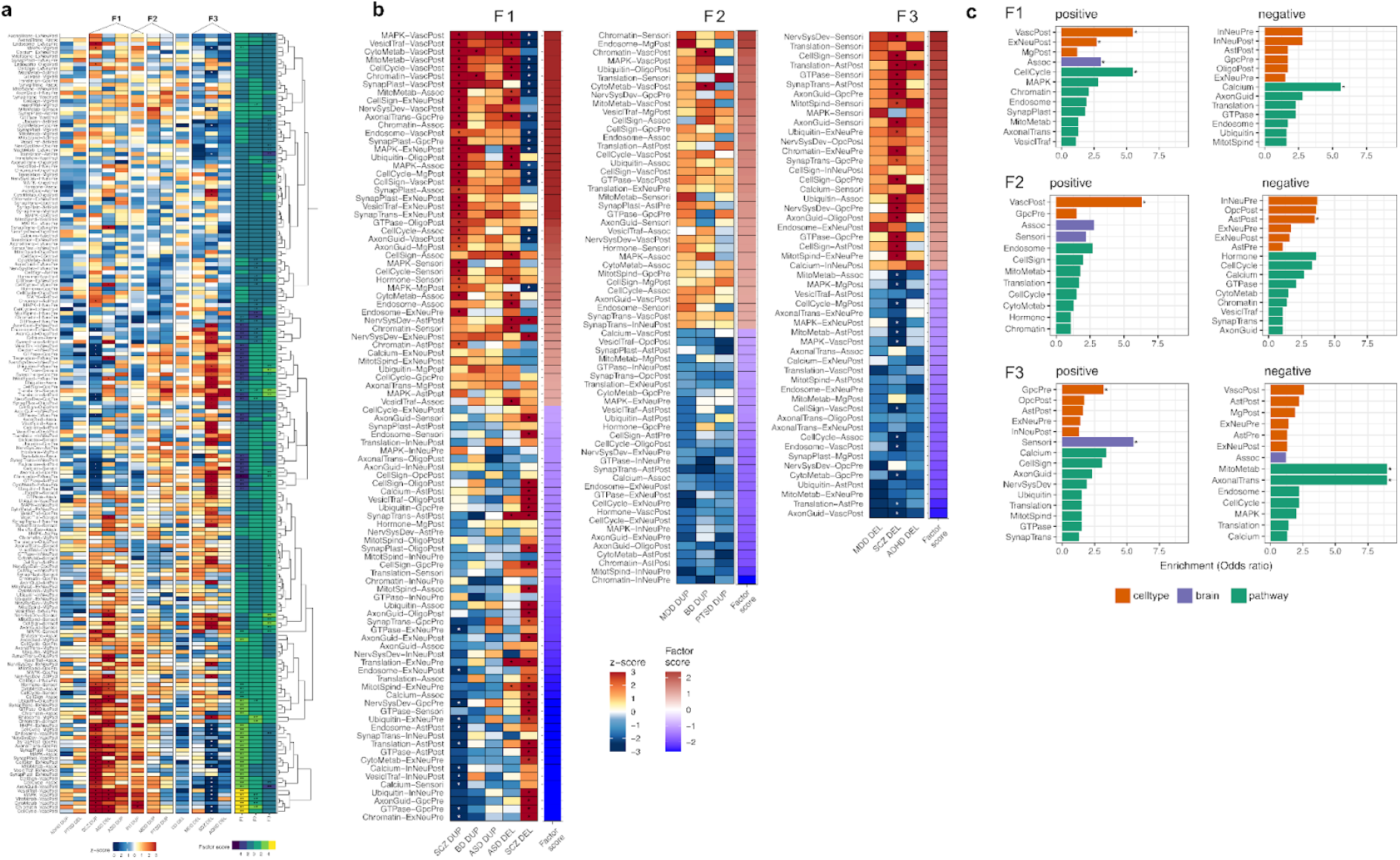
Gene sets and functional terms linked to latent factors F1, F2 and F3 highlight neural processes that underlie orthogonal dimensions of gene-trait relationships. (**a**) a heatmap showing full gene set associations of all two-way pathway-stratified gene-sets (i.e., pathway-cell-type, and pathway-brain stratification). Red-white-blue color scale indicates gene set effect size from sample size weighted meta-analysis (z-score). Yellow-green-blue color scale indicates the F1, F2 and F3 factor scores for each gene set. Asterisks indicate gene set association that meets FDR correction in the combined summary statistics on 6 diagnostic categories (FDR<10%). **factor scores with absolute value >1. (**b**) To illustrate pathway-cell type and pathway-brain associations that contribute to factors, subsets of diagnosis-dosage and gene-sets were selected based on factor loadings and factor scores** for F1, F2 and F3 and sorted by factor score. (**c**) A bar plot highlighting pathway and cell-type terms that were enriched among positively or negatively loaded gene sets in panel B relative to the full summary statistics (fisher exact test P < 0.05).

**Fig. S7:**
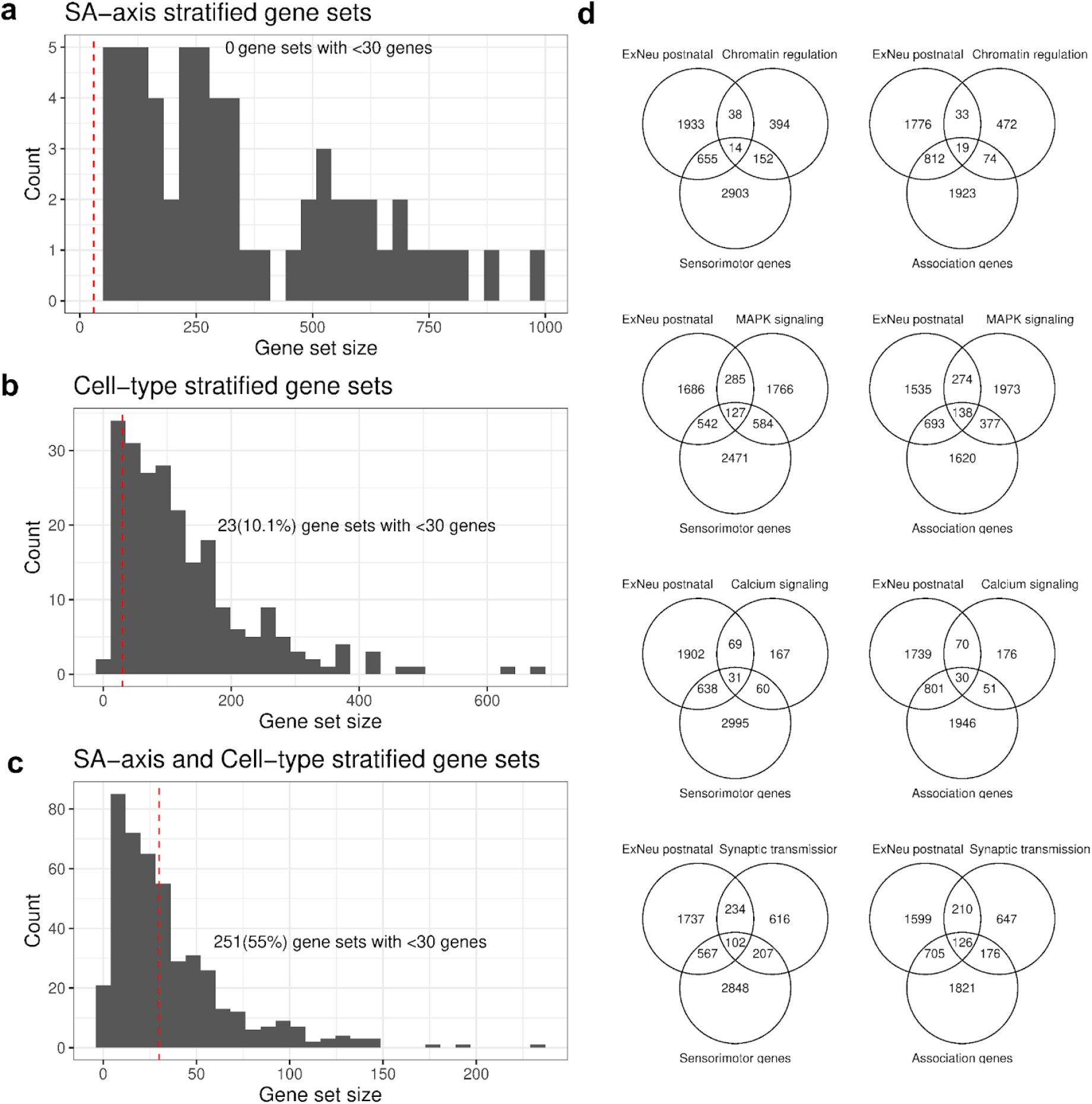
Gene set size of stratified pathways. (**a**)-(**c**) Histograms display the distribution of gene set size when stratified the pathway clusters by (**a**) S-A axis, (**b**) 12 cell types, and (**c**) both S-A axis and cell types. Vertical dashed line indicates our 50 genes cut-off for gene sets to be included in the analysis. (**d**) Venn diagrams show the number of genes intersected between the major pathway gene sets (Chromatin regulation, MAPK signaling, Calcium signaling, and Synaptic transmission), Postnatal Excitatory Neurons, and Sensorimotor or Association genes.

**Fig. S8:**
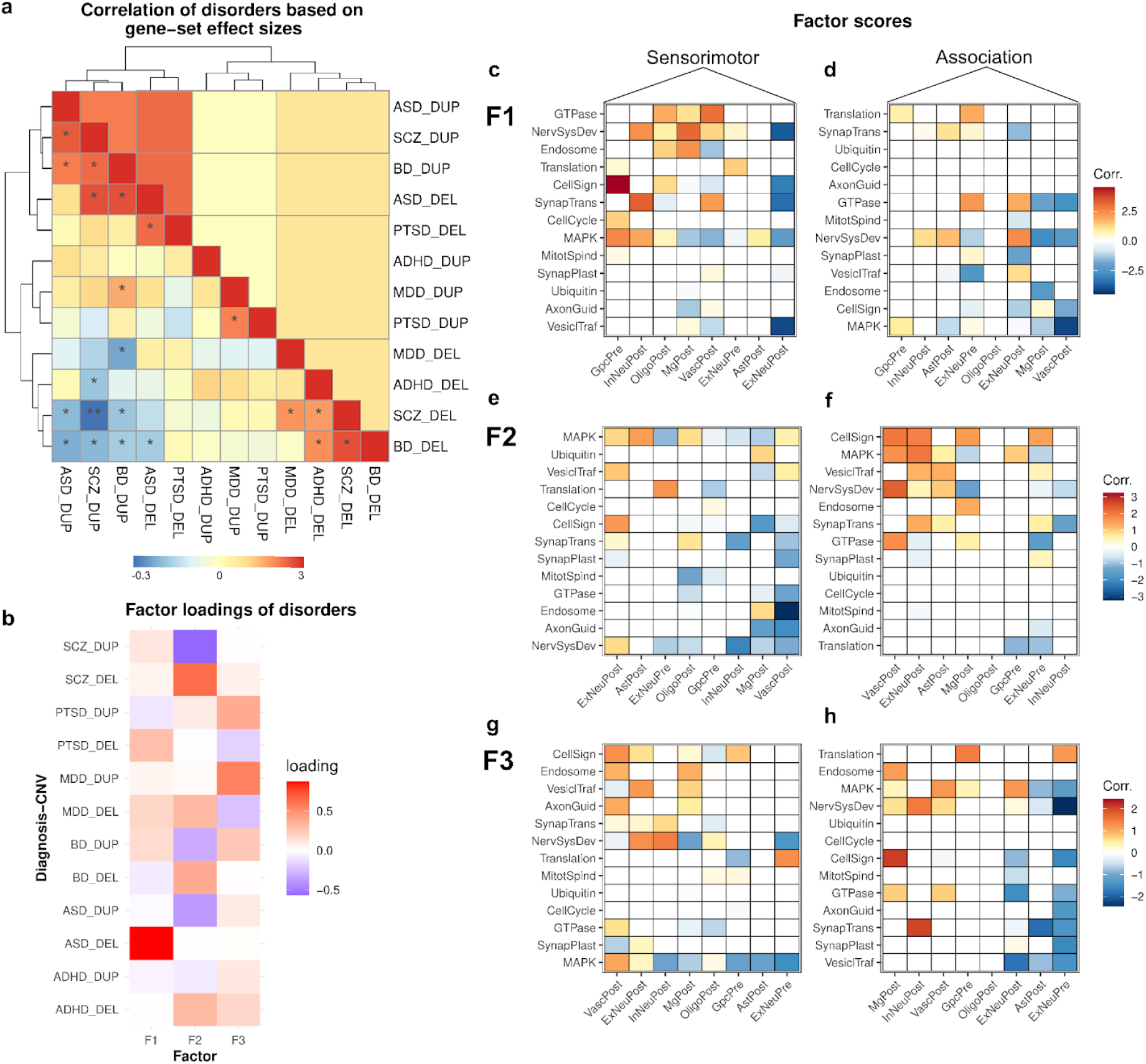
Factor analysis of three-way pathway-celltype-brain stratification. The result shows that factor F2 and F3 are corresponding to the factor F1 and factor F2 of the main factor analysis result (**Fig 5**.) (**a**) Genetic correlation between diagnosis-dosage components. (**b**) Factor loadings. Factor scores for gene sets were shown as heatmaps for each of the three factors; where (**c**) and (**d**) correspond to factor F1, sensorimotor, and association gene sets, respectively. Similarly, (**e**,**f**,**g**,**h**) heatmaps show the factor scores for the factor F2, and F3.

## Notes

### Author Declarations

The original source of the data (i.e., microarray data) used for CNV calling were from other already existed studies prior to this study. All data comes into the PGC de-identified.

### Summary of Updates

This revision is to update the citation to our companionship paper, to standardize the affiliations of the co-authors, and to solve an issue with the supplementary file when open on macOS.

## Reference

1. Parikshak, N. N. et al. Integrative functional genomic analyses implicate specific molecular pathways and circuits in autism. Cell 155, 1008–1021 (2013).

2. Birnbaum, R. & Weinberger, D. R. Genetic insights into the neurodevelopmental origins of schizophrenia. Nat. Rev. Neurosci. 18, 727–740 (2017).

3. Walsh, T. et al. Rare structural variants disrupt multiple genes in neurodevelopmental pathways in schizophrenia. Science 320, 539–543 (2008).

4. McCarthy, S. E. et al. Microduplications of 16p11.2 are associated with schizophrenia. Nat. Genet. 41, 1223–1227 (2009).

5. Marshall, C. R. et al. Contribution of copy number variants to schizophrenia from a genome-wide study of 41,321 subjects. Nat. Genet. 49, 27–35 (2017).

6. Singh, T. et al. Rare coding variants in ten genes confer substantial risk for schizophrenia. Nature 604, 509–516 (2022).

7. Satterstrom, F. K. et al. Large-Scale Exome Sequencing Study Implicates Both Developmental and Functional Changes in the Neurobiology of Autism. Cell 180, 568–584.e23 (2020).

8. Fu, J. M. et al. Rare coding variation provides insight into the genetic architecture and phenotypic context of autism. Nat. Genet. 54, 1320–1331 (2022).

9. Sanders, S. J. et al. Insights into Autism Spectrum Disorder Genomic Architecture and Biology from 71 Risk Loci. Neuron 87, 1215–1233 (2015).

10. Sebat, J. et al. Strong association of de novo copy number mutations with autism. Science 316, 445–449 (2007).

11. Trost, B. et al. Genomic architecture of autism from comprehensive whole-genome sequence annotation. Cell 185, 4409–4427.e18 (2022).

12. Pinto, D. et al. Convergence of genes and cellular pathways dysregulated in autism spectrum disorders. Am. J. Hum. Genet. 94, 677–694 (2014).

13. De Rubeis, S. et al. Synaptic, transcriptional and chromatin genes disrupted in autism. Nature 515, 209–215 (2014).

14. Stahl, E. A. et al. Genome-wide association study identifies 30 loci associated with bipolar disorder. Nat. Genet. 51, 793–803 (2019).

15. Trubetskoy, V. et al. Mapping genomic loci implicates genes and synaptic biology in schizophrenia. Nature 604, 502–508 (2022).

16. Iakoucheva, L. M., Muotri, A. R. & Sebat, J. Getting to the Cores of Autism. Cell 178, 1287–1298 (2019).

17. Singh, T., Neale, B. M. & Daly, M. J. Exome sequencing identifies rare coding variants in 10 genes which confer substantial risk for schizophrenia. MedRxiv (2020).

18. Palmer, D. S. et al. Exome sequencing in bipolar disorder identifies AKAP11 as a risk gene shared with schizophrenia. Nat. Genet. 54, 541–547 (2022).

19. Gandal, M. J. et al. Shared molecular neuropathology across major psychiatric disorders parallels polygenic overlap. Science 359, 693–697 (2018).

20. Gandal, M. J. et al. Transcriptome-wide isoform-level dysregulation in ASD, schizophrenia, and bipolar disorder. Science 362, eaat8127 (2018).

21. Guan, J., Cai, J. J., Ji, G. & Sham, P. C. Commonality in dysregulated expression of gene sets in cortical brains of individuals with autism, schizophrenia, and bipolar disorder. Transl. Psychiatry 9, 152 (2019).

22. Voineagu, I. et al. Transcriptomic analysis of autistic brain reveals convergent molecular pathology. Nature 474, 380–384 (2011).

23. Bowden, N. A., Scott, R. J. & Tooney, P. A. Altered gene expression in the superior temporal gyrus in schizophrenia. BMC Genomics 9, 199 (2008).

24. Velmeshev, D. et al. Single-cell genomics identifies cell type-specific molecular changes in autism. Science 364, 685–689 (2019).

25. Wamsley, B. et al. Molecular cascades and cell type-specific signatures in ASD revealed by single-cell genomics. Science 384, eadh2602 (2024).

26. Rajkowska, G. Cell pathology in bipolar disorder. Bipolar Disord. 4, 105–116 (2002).

27. Skene, N. G. et al. Genetic identification of brain cell types underlying schizophrenia. Nat. Genet. 50, 825–833 (2018).

28. Daskalakis, N. P. et al. Systems biology dissection of PTSD and MDD across brain regions, cell types, and blood. Science 384, eadh3707 (2024).

29. Owen, M. J., Legge, S. E., Rees, E., Walters, J. T. R. & O’Donovan, M. C. Genomic findings in schizophrenia and their implications. Mol. Psychiatry 28, 3638–3647 (2023).

30. Ferreira, M. A. R. et al. Collaborative genome-wide association analysis supports a role for ANK3 and CACNA1C in bipolar disorder. Nat. Genet. 40, 1056–1058 (2008).

31. Verpelli, C. & Sala, C. Molecular and synaptic defects in intellectual disability syndromes. Curr. Opin. Neurobiol. 22, 530–536 (2012).

32. Rosenthal, S. B. et al. A convergent molecular network underlying autism and congenital heart disease. Cell Syst 12, 1094–1107.e6 (2021).

33. Shao, X. et al. Copy number variation is highly correlated with differential gene expression: a pan-cancer study. BMC Med. Genet. 20, 175 (2019).

34. Brunetti-Pierri, N. et al. Recurrent reciprocal 1q21.1 deletions and duplications associated with microcephaly or macrocephaly and developmental and behavioral abnormalities. Nat. Genet. 40, 1466–1471 (2008).

35. Schleifer, C. H. et al. Effects of gene dosage and development on subcortical nuclei volumes in individuals with 22q11.2 copy number variations. Neuropsychopharmacology (2024) doi:10.1038/s41386-024-01832-3.

36. Modenato, C. et al. Effects of eight neuropsychiatric copy number variants on human brain structure. Transl. Psychiatry 11, 399 (2021).

37. Moreau, C. A. et al. Mutations associated with neuropsychiatric conditions delineate functional brain connectivity dimensions contributing to autism and schizophrenia. Nat. Commun. 11, 5272 (2020).

38. Jacquemont, S. et al. Mirror extreme BMI phenotypes associated with gene dosage at the chromosome 16p11.2 locus. Nature 478, 97–102 (2011).

39. Qiu, Y. et al. Oligogenic Effects of 16p11.2 Copy-Number Variation on Craniofacial Development. Cell Rep. 28, 3320–3328.e4 (2019).

40. Shanta, O. et al. A cross-disorder analysis of CNVs finds novel loci and dose-dependent relationships of genes to psychiatric traits. medRxiv 2025.07.11.25331310 (2025).

41. Harris, M. A. et al. The Gene Ontology (GO) database and informatics resource. Nucleic Acids Res. 32, D258–61 (2004).

42. Kanehisa, M., Sato, Y., Kawashima, M., Furumichi, M. & Tanabe, M. KEGG as a reference resource for gene and protein annotation. Nucleic Acids Res. 44, D457–62 (2016).

43. Fabregat, A. et al. Reactome pathway analysis: a high-performance in-memory approach. BMC Bioinformatics 18, 142 (2017).

44. Velmeshev, D. et al. Single-cell analysis of prenatal and postnatal human cortical development. Science 382, eadf0834 (2023).

45. Glasser, M. F. et al. A multi-modal parcellation of human cerebral cortex. Nature 536, 171–178 (2016).

46. Hawrylycz, M. J. et al. An anatomically comprehensive atlas of the adult human brain transcriptome. Nature 489, 391–399 (2012).

47. Subramanian, A. et al. Gene set enrichment analysis: a knowledge-based approach for interpreting genome-wide expression profiles. Proc. Natl. Acad. Sci. U. S. A. 102, 15545–15550 (2005).

48. Nishimura, D. BioCarta. Biotech Softw. Internet Rep. 2, 117–120 (2001).

49. Merico, D., Isserlin, R., Stueker, O., Emili, A. & Bader, G. D. Enrichment map: a network-based method for gene-set enrichment visualization and interpretation. PLoS One 5, e13984 (2010).

50. Antaki, D. et al. A phenotypic spectrum of autism is attributable to the combined effects of rare variants, polygenic risk and sex. Nat. Genet. 54, 1284–1292 (2022).

51. Dear, R. et al. Cortical gene expression architecture links healthy neurodevelopment to the imaging, transcriptomics and genetics of autism and schizophrenia. Nat. Neurosci. 27, 1075–1086 (2024).

52. Markello, R. D. et al. Neuromaps: Structural and functional interpretation of brain maps. Nat. Methods 19, 1472–1479 (2022).

53. Misic, B. et al. Neuromaps: structural and functional interpretation of brain maps. Research Square (2022) doi:10.21203/rs.3.rs-1296849/v1.

54. Paus, T., Keshavan, M. & Giedd, J. N. Why do many psychiatric disorders emerge during adolescence? Nat. Rev. Neurosci. 9, 947–957 (2008).

55. Burt, J. B. et al. Hierarchy of transcriptomic specialization across human cortex captured by structural neuroimaging topography. Nat. Neurosci. 21, 1251–1259 (2018).

56. Sydnor, V. J. et al. Neurodevelopment of the association cortices: Patterns, mechanisms, and implications for psychopathology. Neuron 109, 2820–2846 (2021).

57. Hilgetag, C. C. & Goulas, A. ‘Hierarchy’ in the organization of brain networks. Philos. Trans. R. Soc. Lond. B Biol. Sci. 375, 20190319 (2020).

58. Mesulam, M. M. From sensation to cognition. Brain 121 (Pt 6), 1013–1052 (1998).

59. Alexander-Bloch, A. F. et al. On testing for spatial correspondence between maps of human brain structure and function. Neuroimage 178, 540–551 (2018).

60. Luo, L., Arizmendi, C. & Gates, K. M. Exploratory factor analysis (EFA) programs in R. Struct. Equ. Modeling 26, 819–826 (2019).

61. Karayiorgou, M. & Gogos, J. A. The molecular genetics of the 22q11-associated schizophrenia. Brain Res. Mol. Brain Res. 132, 95–104 (2004).

62. Kopp, N., McCullough, K., Maloney, S. E. & Dougherty, J. D. Gtf2i and Gtf2ird1 mutation do not account for the full phenotypic effect of the Williams syndrome critical region in mouse models. Hum. Mol. Genet. 28, 3443–3465 (2019).

63. Sefik, E. et al. Convergent and distributed effects of the 3q29 deletion on the human neural transcriptome. Transl. Psychiatry 11, 357 (2021).

64. Grotzinger, A. D. et al. The landscape of shared and divergent genetic influences across 14 psychiatric disorders. medRxiv 2025.01.14.25320574 (2025) doi:10.1101/2025.01.14.25320574.

65. So, H.-C., Chau, K.-L., Ao, F.-K., Mo, C.-H. & Sham, P.-C. Exploring shared genetic bases and causal relationships of schizophrenia and bipolar disorder with 28 cardiovascular and metabolic traits. Psychol. Med. 49, 1286–1298 (2019).

66. Veeneman, R. R. et al. Exploring the relationship between schizophrenia and cardiovascular disease: A genetic correlation and multivariable Mendelian randomization study. Schizophr. Bull. 48, 463–473 (2022).

67. Correll, C. U. et al. Prevalence, incidence and mortality from cardiovascular disease in patients with pooled and specific severe mental illness: a large-scale meta-analysis of 3,211,768 patients and 113,383,368 controls. World Psychiatry 16, 163–180 (2017).

68. Fagiolini, A. & Swartz, H. A. Cardiovascular disease and bipolar disorder: Risk and clinical implications. Psychiatrist.com (2012).

69. Dhanasekara, C. S. et al. Association between autism spectrum disorders and cardiometabolic diseases: A systematic review and meta-analysis: A systematic review and meta-analysis. JAMA Pediatr. 177, 248–257 (2023).

70. Canitano, R. & Pallagrosi, M. Autism spectrum disorders and schizophrenia spectrum disorders: Excitation/inhibition imbalance and developmental trajectories. Front. Psychiatry 8, 69 (2017).

71. Dzafic, I. et al. Stronger top-down and weaker bottom-up frontotemporal connections during sensory learning are associated with severity of psychotic phenomena. Schizophr. Bull. 47, 1039–1047 (2021).

72. Lyu, X. et al. Weaker top-down cognitive control and stronger bottom-up signaling transmission as a pathogenesis of schizophrenia. Schizophrenia (Heidelb.) 11, 36 (2025).

73. Karayiorgou, M., Simon, T. J. & Gogos, J. A. 22q11.2 microdeletions: linking DNA structural variation to brain dysfunction and schizophrenia. Nat. Rev. Neurosci. 11, 402–416 (2010).

74. Jacquemont, S. et al. Genes To Mental Health (G2MH): A Framework to Map the Combined Effects of Rare and Common Variants on Dimensions of Cognition and Psychopathology. Am. J. Psychiatry 179, 189–203 (2022).

75. Urresti, J. et al. Cortical organoids model early brain development disrupted by 16p11.2 copy number variants in autism. Mol. Psychiatry 26, 7560–7580 (2021).

76. Nehme, R. et al. The 22q11.2 region regulates presynaptic gene-products linked to schizophrenia. Nat. Commun. 13, 3690 (2022).

77. Kumar, K. et al. Cortical differences across psychiatric disorders and associated common and rare genetic variants. medRxiv 2025.04.16.25325971 (2025) doi:10.1101/2025.04.16.25325971.

78. Abraham, G., Qiu, Y. & Inouye, M. FlashPCA2: principal component analysis of Biobank-scale genotype datasets. Bioinformatics 33, 2776–2778 (2017).

79. Chen, C.-Y. et al. Improved ancestry inference using weights from external reference panels. Bioinformatics 29, 1399–1406 (2013).

80. Nievergelt, C. M. et al. International meta-analysis of PTSD genome-wide association studies identifies sex- and ancestry-specific genetic risk loci. Nat. Commun. 10, 4558 (2019).

81. Das, S. et al. Late onset psychosis in a case of 15q11.2 BP1-BP2 microdeletion (Burnside-Butler) syndrome: A case report and literature review. SAGE Open Med. Case Rep. 12, 2050313×241229058 (2024).

82. Markello, R. D. et al. Standardizing workflows in imaging transcriptomics with the abagen toolbox. bioRxiv (2021) doi:10.1101/2021.07.08.451635.

83. Kumar, K. et al. Mirror effect of genomic deletions and duplications on cognitive ability across the human cerebral cortex. bioRxiv 2025.01. 06.631492 (2025) doi:10.1101/2025.01.06.631492.

84. Mowinckel, A. M. & Vidal-Piñeiro, D. Visualization of brain statistics with R packages ggseg and ggseg3d. Adv. Methods Pract. Psychol. Sci. 3, 466–483 (2020).

